# Effect of a 24-week resistance exercise intervention on cognitive function in cognitively normal older adults: The AGUEDA Randomized Controlled Trial

**DOI:** 10.1101/2025.04.29.25326693

**Authors:** Beatriz Fernandez-Gamez, Patricio Solis-Urra, Andrea Coca-Pulido, Cristina Molina-Hidalgo, Marcos Olvera-Rojas, Esmée A. Bakker, Darío Bellón, Alessandro Sclafani, Jose Mora-Gonzalez, Javier Fernández-Ortega, Lucía Sánchez-Aranda, Isabel Martín-Fuentes, Angel Toval, Javier Sanchez-Martinez, Lu Wan, Manuel Gomez-Rio, Teresa Liu-Ambrose, Kirk I. Erickson, Francisco B. Ortega, Irene Esteban-Cornejo

## Abstract

**Objective:** The AGUEDA trial examined the effects of a 24-week resistance exercise (RE) intervention on executive function (EF) and other cognitive domains in cognitively normal older adults.

**Method:** Ninety participants (71.75 ± 3.96 years, 57.8% female) were randomized to an RE group (n = 46) or a control group (n = 44). The RE group performed 180 minutes/week of supervised training, while the control group maintained usual activities. EF and other cognitive domains (e.g., attentional/inhibitory control, episodic memory, processing speed, visuospatial processing, and working memory) were assessed at baseline, and 24 weeks. Intervention effects were analyzed using intention-to-treat (ITT) and per-protocol (PP) approaches, with exploratory subgroup analyses based on sociodemographic and AD-related characteristics.

**Results:** EF composite score improved in both groups, with no significant between-group differences (standardized mean difference (SMD) = 0.13, p = 0.37). In addition, the RE group showed significant improvements in attentional/inhibitory control compared to the control group (SMD = 0.43, p < 0.001), while the rest of cognitive domains showed no significant differences (p > 0.05). Subgroup analyses revealed greater EF benefits for the oldest adults, those with lower educational levels, and individuals with higher subjective cognitive decline (SCD) at baseline. RE had an effect on knee extension strength (SMD = 0.25, p = 0.02), which was positively correlated with better EF (r = 0.38, p = 0.0005) and improved episodic memory (r = 0.31, p = 0.008).

**Conclusion:** The 24-week AGUEDA intervention revealed no significant differences in EF or other cognitive domains after 24 weeks. However, participants in the RE group demonstrated significantly greater improvements in attentional/inhibitory control compared to the control group. Moreover, our findings support the notion that RE can yield greater benefits in the more vulnerable subgroups, such as the oldest participants, those with SCD or fewer years of education. Although the mediation analysis did not find significant indirect effects, lower body muscular strength improvements were positively correlated with better EF and episodic memory, highlighting the potential role of strength in cognitive health.

## 1. Introduction

Dementia is a major cause of disability worldwide (1–3) making it one of today’s most pressing public health challenges. The absence of treatments to slow down disease progression that leads to dementia underscores the preventive approaches for protection of cognitive function (3–6). Exercise is one of the most promising non-pharmaceutical interventions to maintain and/or enhance cognitive function in late life (4,5,7). While most of the research has focused on aerobic exercise training (8–14) (i.e., exercise that involves continuous movements of the body’s large muscles in a rhythmic manner for sustained periods (15)), recent meta-analyses suggested that resistance exercise (RE) could have valuable beneficial effects on cognitive domains compared to other exercise modalities (16,17). In fact, it seems like RE at lower doses may be already enough to achieve clinically meaningful changes (4).

RE using elastic bands and body weight may be an accessible and economic alternative to traditional free-weight or machine-based interventions, with focus on health benefits (18–20). Prior exercise interventions on the cognitive effects of elastic-band and body-weight RE has primarily relied on global cognitive screening measures (e.g., Montreal Cognitive Assessment (MoCA), Mini-Mental State Examination (MMSE)) (21,22), or single cognitive test to measure a domain (e.g., Rey Auditory Verbal Learning Test for episodic memory or Trail Making Test for attention (23)) with fewer studies employing specific tests targeting distinct cognitive domains (e.g., executive function (EF) (23), attention (24), processing speed (25) or memory (26)). In this context, recent findings suggest that identifying the cognitive domains most responsive to exercise-mediated improvements could facilitate the detection of individuals who are more likely to benefit from such interventions (27). Hence, it is still unclear whether RE, particularly when performed with elastic bands, exerts broad effects across global cognitive function or if certain cognitive domains derive the greatest benefit (28).

Individual demographic or genetic characteristics, such as Apolipoprotein E ε4 (APOE4) (29), age (30) or Subjective Cognitive Decline (SCD) (31) may also moderate the effects of RE on cognition in older adults (32). Previous studies with aerobic exercise have reported greater cognitive effect in males (33), APOE4 carriers (34) or in those with high educational level (35). Interestingly, studies with RE interventions found greater cognitive benefits in females (36), APOE4 noncarriers (37), in oldest participants (38), those with lower educational levels (39) or those with higher SCD (40). Thus, these findings underscore the complexity of understanding the role of individuals characteristics in moderating the cognitive effects of exercise, and specifically of RE (41).

Well-designed randomized controlled trials (RCTs) examining the effects of easily prescribed RE interventions, along with the role of potential moderators, are essential for advancing our understanding of how to optimize cognitive function in aging. These findings could contribute to public health guidelines by providing tailored exercise recommendations for preserving cognitive health in older adults. The AGUEDA trial was designed to investigate the effects of a 24-week RE intervention using elastic-bands and body-weight on cognitive function in cognitively normal older adults. The primary outcome was an EF composite score, while secondary outcomes included other cognitive domains (i.e., attentional/inhibitory control, episodic memory, processing speed, visuospatial processing, and working memory). Secondly, we examined the effects of several individual-level moderators (i.e., sex, age, educational level, comorbidities, APOE4 carriership, amyloid burden, baseline cognitive performance, and baseline SCD). Thirdly, we explored the e ects of RE on physical condition parameters. (i.e., muscular strength, physical function and cardiorespiratory fitness). Finally, we explored whether any of these exercise-related outcomes statistically mediated the exercise-derived cognitive improvements. The overall hypothesis was that a 24-week RE intervention would improve selective cognitive domains in cognitively normal older adults, particularly in the more vulnerable subgroups, (e.g., the oldest participants, those with SCD or with fewer years of education), and these cognitive benefits will be mediated by improvements in muscular strength.

## 2. Methods

### 2.1 Design and participants

The AGUEDA trial was a single-center, two-arm, single-blind RCT that randomized 90 cognitively normal older adults (65-80 years old) from Granada (Spain). All data were collected between March, 2021 and May, 2022. The trial protocol followed the principles of the Declaration of Helsinki and was approved by the Research Ethics Board of the Andalusian Health Service (CEIM/CEI Provincial de Granada; #2317-N-19) on May 25th, 2020. The trial was registered on Clinicaltrials.gov (Clinicaltrials.gov Identifier: NCT05186090; Submission date: December 22, 2021). All participants provided written informed consent. The reporting of the results adheres to the Consolidated Standards of Reporting Trials Extension (CONSORT extension) guidelines (42) (**Supplementary material 1, Table S1).** All outcome-related measures and analyses are performed by sta who were blinded to the intervention assignment. Details of the methodology and protocols of the AGUEDA project have been described elsewhere (43) and deposited at GitHub (https://github.com/aguedaprojectugr).

### 2.2 Eligibility criteria

Eligibility criteria were defined as: (i) older adults between 65 - 80 years old, (ii) physically inactive (i.e., defined as not participating in any RE program in the last 6 months or accumulating less than 600 METs-Min/week by the International Physical Activity Questionnaire (IPAQ)) (44), (iii) cognitively normal (≥ 26 points) according to the Spanish version of the modified Telephone Interview of Cognitive Status (STICS-m) (45), MMSE (≥ 25/30) (46) and MoCA (<71 years: ≥24/30; 71-75 years: 22/30; >75 years: 21/30) (47); and (iv) not to present significant depressive symptoms at baseline according to the Geriatric Depression Scale (GDS) (≥15) (48). Detailed information about eligibility criteria is detailed in **Supplementary material 2, Table S3,** and available elsewhere (43).

### 2.3 Intervention and control

Details of the methodology of the AGUEDA exercise intervention have been fully described elsewhere following the Consensus on Exercise Reporting Template (CERT) (49). Briefly, participants randomly assigned to the 24-week RE intervention group were asked to attend 3 sessions per week, for 60 min each day (10 min for the warm-up phase, 45 min for the main phase, and 5 min for the cool-down phase) at the Sport and Health University Research Institute research center (iMUDS), in Granada, Spain. The RE intervention consisted of a combination of upper and lower limb exercises using elastic bands and the participant’s body weight. The volume and intensity were based on the resistance of the elastic bands (TheraBand; 7 resistances divided by colors) (50), number of repetitions (individualized), motor complexity of exercises (3 levels), sets and rest (3 sets/60-sec rest), execution time (40-60 sec) and velocity (as fast as possible). The control group was asked to maintain their usual lifestyle. After the 24-week period, they were offered the RE intervention for ethical reasons. The intervention description follows the Template for Intervention Description and Replication (TIDieR) guidelines (51) (Supplementary material 1, Table S2).

### 2.4 Power calculation and sample size

We based the power calculation on a meta-analysis showing that the effect size of exercise interventions on EF composite score in older adults was 0.34 (95% Confidence interval = 0.22 to 0.47) (52) with a two-tailed alpha at 0.05 and a power of 80%. After adjusting for a 20% estimated dropout rate, 45 participants for each group were needed for sufficient power.

### 2.5 Outcomes measurements

The primary outcome (EF composite score) was assessed at baseline, at mid-point (12 weeks), and post-intervention (24 weeks), while secondary and other outcomes were assessed only at baseline and post-intervention. Information about the outcome’s measurements are detailed in **Supplementary material 3, 4 and 5.**

#### 2.5.1 Primary outcome: Executive function

An EF composite score was created by performing confirmatory factor analyses (CFA). The cognitive tests included in the CFA were pre-specified in the methodology of the AGUEDA trial (43) based on available evidence and theoretical positions of EF (53). A first-order factor CFA was performed for the final EF composite score which the following cognitive indicators: Trail Making Test (TMT) (Interference Score, Time part B – Time part A) (54), Digit Symbol Substitution test (DSST) (Total correct score) (55), Dimensional Change Card Sort Test (DCCST) (Inverse efficiency score, reaction time/accuracy) (56) and Spatial Working Memory Test (SWMT) (Inverse efficiency score of switch trials – high load, reaction time/accuracy) (57).

Other tests were included in the first-order factor CFA but were not retained in the final EF composite score (i.e., Picture Sequence Memory Test (PSMT) (56), List Sorting Working Memory Test (LSWMT) (56), Flanker test (56), Stroop test, and Task switching test (58)). All methodological details and results of the CFA are provided in **Supplementary material 3, Section 2**.

#### 3.5.2 Secondary outcomes: Other cognitive domains– attentional/inhibitory control, episodic memory, processing speed, visuospatial processing, and working memory

Participants completed a comprehensive neuropsychological evaluation that measured different cognitive domains detailed in **Supplementary material 3, Section 1, 3 and Table S8**:

i. Attentional/inhibitory control: TMT (Time part B) (54), DCCST (Computed score) (59), Stroop test (Incongruent raw score), Flanker test (Computed score) (59).
ii. Episodic memory: MoCA delayed recall (Multiple choice cue) (60), PSMT (Raw score) (59), Rey Auditory Verbal Learning Test (Total recall, delayed and recognition raw score) (61), Rey - Osterrieth Complex Figure Test (ROFT) (Copy and delayed raw score) (62).
iii. Processing speed: DSST (Total correct raw score) (55) and TMT, (Time part A) (54).
iv. Visuospatial processing: Wechsler Adult Intelligence Scale with Matrix Reasoning and Block Design (Accuracy) (63) and MoCA Clock Draw (Total Score) (60).
v. Working memory: SWMT (3 & 4 items) (64), LSWMT (Total correct) (65), N-Back Test (2-back) (66).

#### 2.5.3 Moderation outcomes

Sociodemographic and AD-related related variables were examined as potential moderators, as detailed in **Supplementary material 4.** These included: (i) sex assigned at birth (male vs female), (ii) age (< 72 years old vs ≥ 72 years), (iii) education (< 12 years vs ≥ 12 years of education), (iv) number of comorbidities (< 3 vs ≥ 3-4), (v) APOE4 carrier (carrier vs non-carrier), (vi) amyloid burden (positive vs negative, 12 CL), (vi) baseline cognitive performance of each domain (< median vs ≥ median) and (vii) median baseline SCD (< 3 points vs ≥ 3 points).

#### 2.5.4 Physical condition parameters.: Muscular strength, physical function and cardiorespiratory fitness

Participants completed a set of physical tests to assess muscular strength, physical function, and cardiorespiratory fitness. Muscular strength, assessed for measurements included both upper and lower body muscular strength. Upper body muscular strength was gauged through the arm curl test from Senior Fitness Test (67), hand dynamometer (TKK 5101 Grip D, Takey, Tokyo, Japan), and the elbow extension strength test by the Gymmex Iso-2 dynamometer (EASYTRCH s.r.l., Italy)(68). Lower body strength was evaluated using the 30-seconds sit-to-stand test from SFT (67), the 5-times sit-to-stand test from Short Physical Performance Battery (SPPB) (69), and the knee extension strength test by the Gymmex Iso-2 dynamometer (EASYTRCH s.r.l., Italy) (70,71). Arm Curl test was measured in the dominant arm, and for both the hand dynamometer and the Gymmex Iso-2 dynamometer, measurements were taken as the average of both limbs. Physical function was assessed by the 2-minute step test and the up and go test, from the SFT (72). In addition, cardiorespiratory fitness was assessed by the 6-minute walk test (73) and the 2-km walking test (74). All methodological details are provided in **Supplementary material 5.**

### 2.6 Intervention attendance and compliance

Session attendance was calculated as the proportion of sessions completed out of the total prescribed sessions (i.e., 72 sessions scheduled over 24 weeks at a frequency of three sessions per week). 80% attendance was required to be included for the per-protocol (PP) analysis (i.e., > 57 exercise sessions). Any missed session was registered and rescheduled to an alternative date to maximize intervention attendance. Compliance was measured by 10-point Borg Rating of Perceived Exertion scale (RPE) (75) with a target rating from 4 to 8 depending on the prescribed intensity level for each week.

### 2.7 Adverse events

All adverse events were reviewed and recorded in REDCap (76), preferably at the time of the event, and additionally asked at the midpoint and post-intervention assessments (i.e., at 12 and 24 weeks, respectively) via phone call to participants in both groups (e.g., RE and control group). The severity of adverse events was classified into 3 categories (i.e., mild, moderate, severe) (77).

### 2.8 Statistical analyses

#### 2.8.1 Main analysis

We conducted main analyses based on both available-case intention-to-treat (ITT), which included all randomized participants, and PP, including participants attending at least 80% of sessions (> 57 sessions).

Data preprocessing involved reviewing the biological plausibility of the data to identify implausible values resulting from suboptimal testing conditions. The 12-week and 24-week outcomes z-scores were calculated, using the baseline mean and SD for each variable. The use of these z score values in whole sample analyses allows comparisons across outcomes of different nature and the z score of change (i.e., post-intervention z score) can be interpreted as standardized effect size as it indicates how many SDs the outcomes at post-intervention have changed with respect to the baseline mean and SD (78). For sensitivity analyses, winsorization was applied to the raw scores of each outcome, when necessary, to reduce the influence of extreme values (i.e., scores ±4SD from the mean). Extreme values were replaced with the nearest value within the acceptable range (78)

Intervention effects were evaluated by examining between-group differences (exercise versus control) from baseline to 24-weeks in EF, other cognitive domains (i.e., attentional/inhibitory control, episodic memory, processing speed, visuospatial processing and working memory), and physical exercise related outcomes (i.e., muscular strength, physical function and cardiorespiratory fitness) following Constrained Linear Mixed Models (CLMM) (i.e., baseline adjusted), specifically with “*LMMstar”* and *“Lmer”* packages in R Studio (79), with restricted maximum likelihood estimation. The models included time and group as a categorical fixed effect, and group-by-time interaction, with the intercept specified as a random effect. Unequal variance was allowed across time and group. Statistical significance was set at p < 0.05 without adjustments for multiple comparisons, as the primary analysis was based on baseline and 24-week comparisons. Estimated marginal means, within-group differences, and between-group differences were calculated by “*emmeans”* package. The intervention effect was represented by the coefficient for the interaction term in the model, along with its (95% confidence intervals (CIs)). Analyses were carried out using R Studio, version 6.1 (R Project for Statistical Computing).

#### 2.8.2 Subgroup analysis

Interaction terms were included and stratified analyses were performed to examine whether key variables (i.e., sex, age, educational level, comorbidities, APOE4 carriership, amyloid burden, baseline cognitive performance, and SCD) modified the effect of the intervention on EF and other cognitive domains. The models followed the same statistical approach as in the primary analysis, employing CLMM, specifically using the “*LMMstar”* and “*lme4”* packages (79) with restricted maximum likelihood estimation. All interactions were interpreted as p < 0.1.

#### 2.8.2 Post hoc mediation analysis

Mediation analyses following AGReMA (A Guideline for Reporting Mediation Analyses) were conducted using the “*lme4”* and “*mediation”* packages in R (79). Muscular strength, physical function and cardiorespiratory fitness indicators were proposed as potential mediators of the effects of exercise on EF and other cognitive domains. Only the indicators showing significant effects were assessed to understand the mechanisms through which the intervention could exert its effect. Standardized (β) regression coefficients are presented for four equations: Equation 1 regressed the mediator (e.g., change in knee extension) on the independent variable (exercise vs control). Equation 2 regressed the dependent variables (i.e., cognitive domain) on the independent variable. Equation 3 regressed the dependent variables on both the mediator (equation 3) and the independent variable (equation 3’). We included the outcome of interest at baseline as a covariate. The indirect effects, along with their 95% CIs are presented, and the significance is considered if the indirect effect significantly differed from zero (i.e., zero was not contained within the CIs). The percentage of the total effect was computed to know how much of the total effect was explained by the mediation, as follows: (indirect effect / total effect) × 100.

#### 2.8.3 Adverse event

A sensitivity analysis was performed using an ITT principle with all randomly assigned participants, but participants who encountered adverse events, which research has shown could impact the primary outcomes (e.g., stroke (80) and cancer (81)) were excluded.

## 3. Results

### 3.1 Sample characteristics

A total of 289 participants were assessed for eligibility by phone call. Of these, 183 provided consented to participate and 90 participants were randomized (flowchart with specific numbers and reason of exclusion is shown in **Figure 1**). Participants demographic characteristics are shown in **Supplementary material 6, Table S10**. The mean age at baseline was 71.75 ± 3.96 years and 52 (57.8%) females were included in the study. The mean level of education was 11.54 ± 4.90 years. Among the participants, 13 (14.8%) were classified as APOE4 carriers, and 19 (21.1%) as positive amyloid burden. Additionally, 37 participants (41.1%) had more than two comorbidities, and the mean score for SCD was 2.92 ± 1.97.

**Figure 1:**
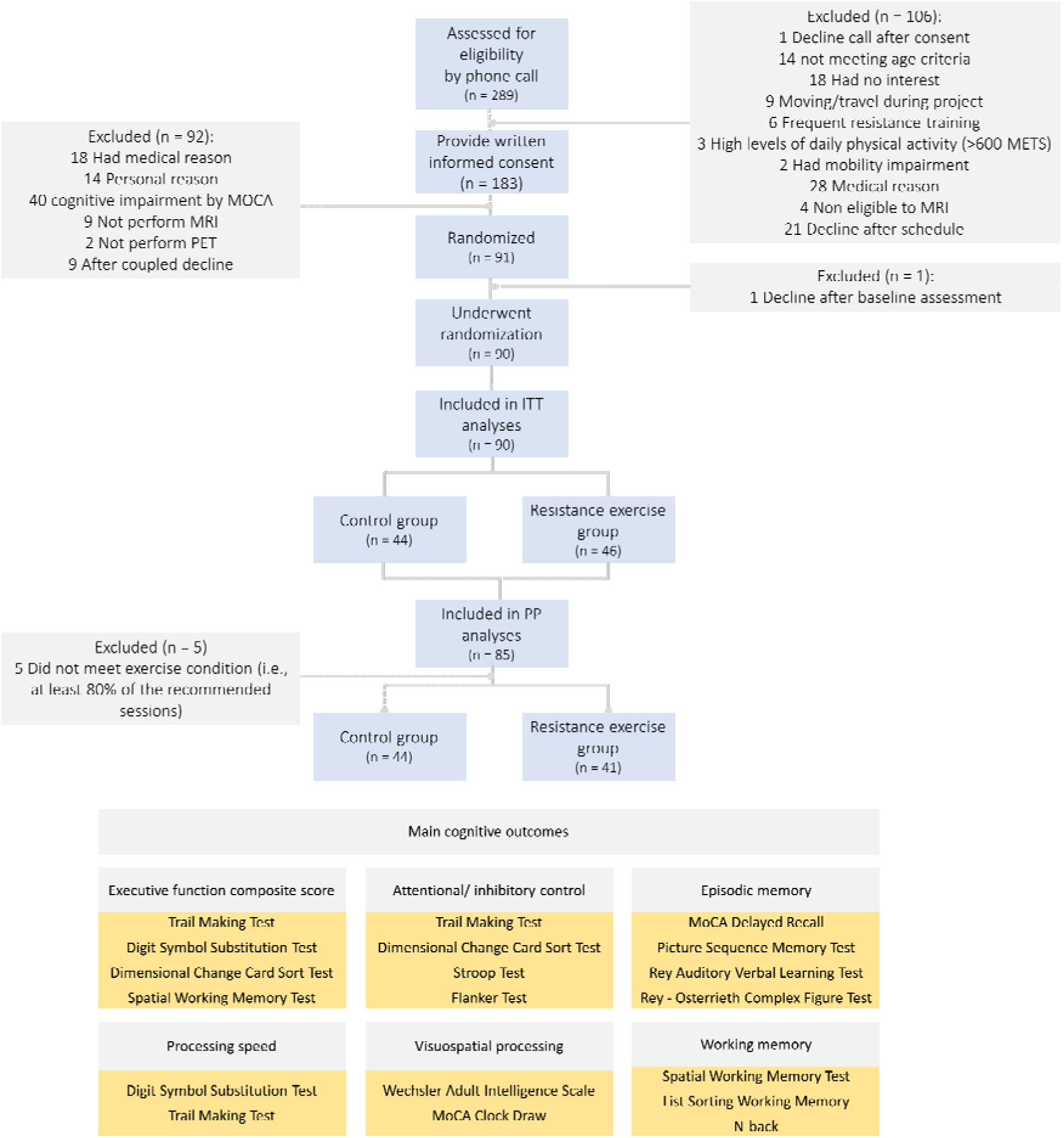
CONSORT Flow diagram of the AGUEDA trial.

### 3.2 Primary outcome

Both exercise and control groups showed significant improvements on the EF composite score at 24 weeks, but we did not detect differences between groups (SMD = 0.13, p = 0.37). In the exercise group, the post intervention change was 0.39 (95% CI, 0.14; 0.65), while in the control group, the change was 0.26 (95% CI, −0.01; 0.53) (**Figure 2A, 2B**).

**Figure 2:**
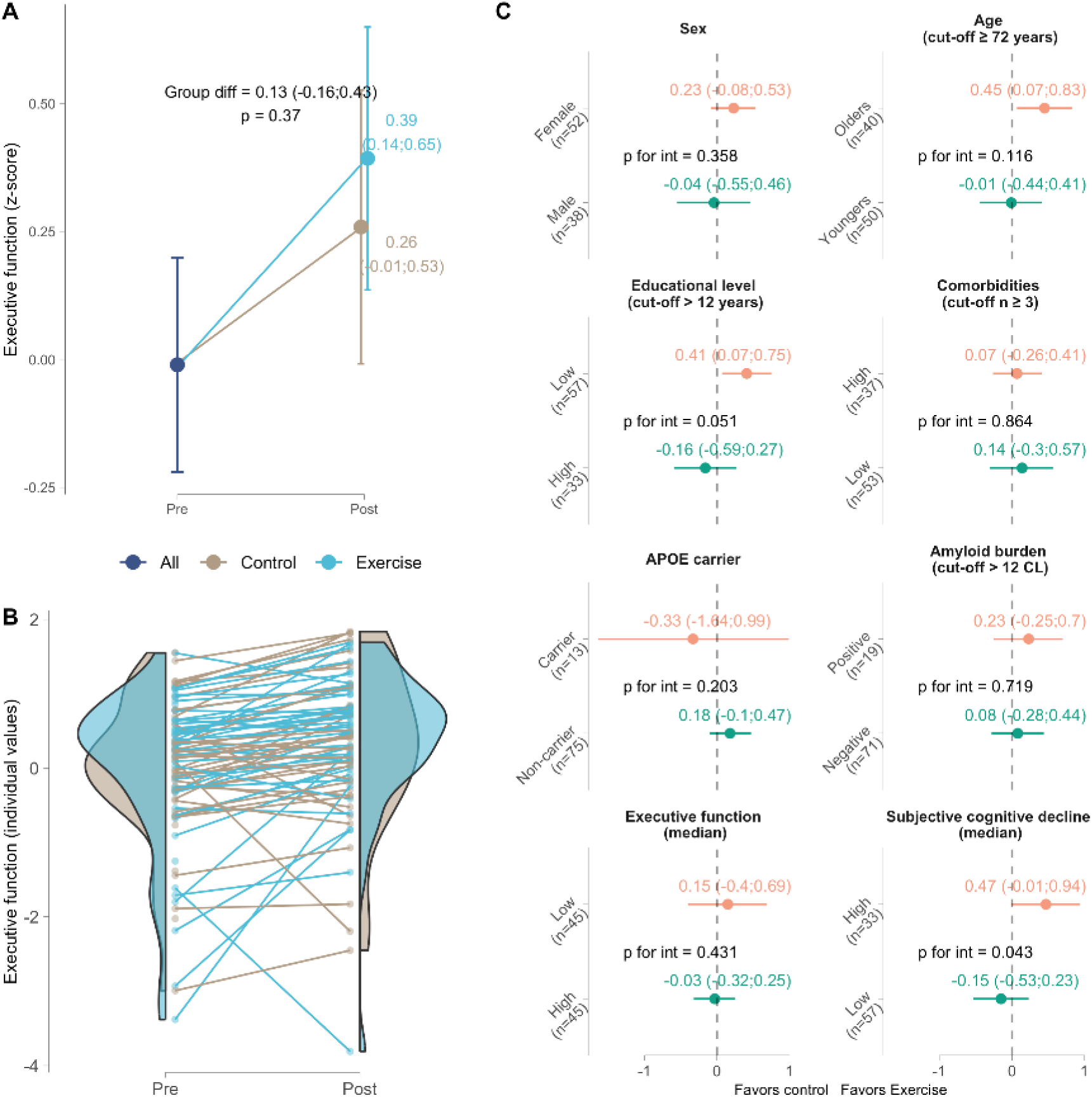
Intention-to-treat effects of the 24-week resistance exercise intervention on executive function for the whole sample (A–mean values; B–individual values) and for subgroups (C). A. Dots indicate estimated marginal means at each time point (and 95% Confidence Intervals). B. Dots indicate participant individual raw values. C. Dots indicate mean difference of marginal means and 95% Confidence Intervals) between exercise and control in each subgroup.

### 3.3 Secondary outcomes

The RE group reported significantly better attentional/inhibitory control than the control group at 24 weeks (SMD = 0.43, p < 0.001). Additionally, no differences were detected between groups in changes on episodic memory (SMD = 0.29, p = 0.12), processing speed (SMD = −0.04, p = 0.77), visuospatial processing (SMD = 0.07, p = 0.71) or working memory (SMD = 0.09, p = 0.60) at 24 weeks (**Figure 3A**).

**Figure 3:**
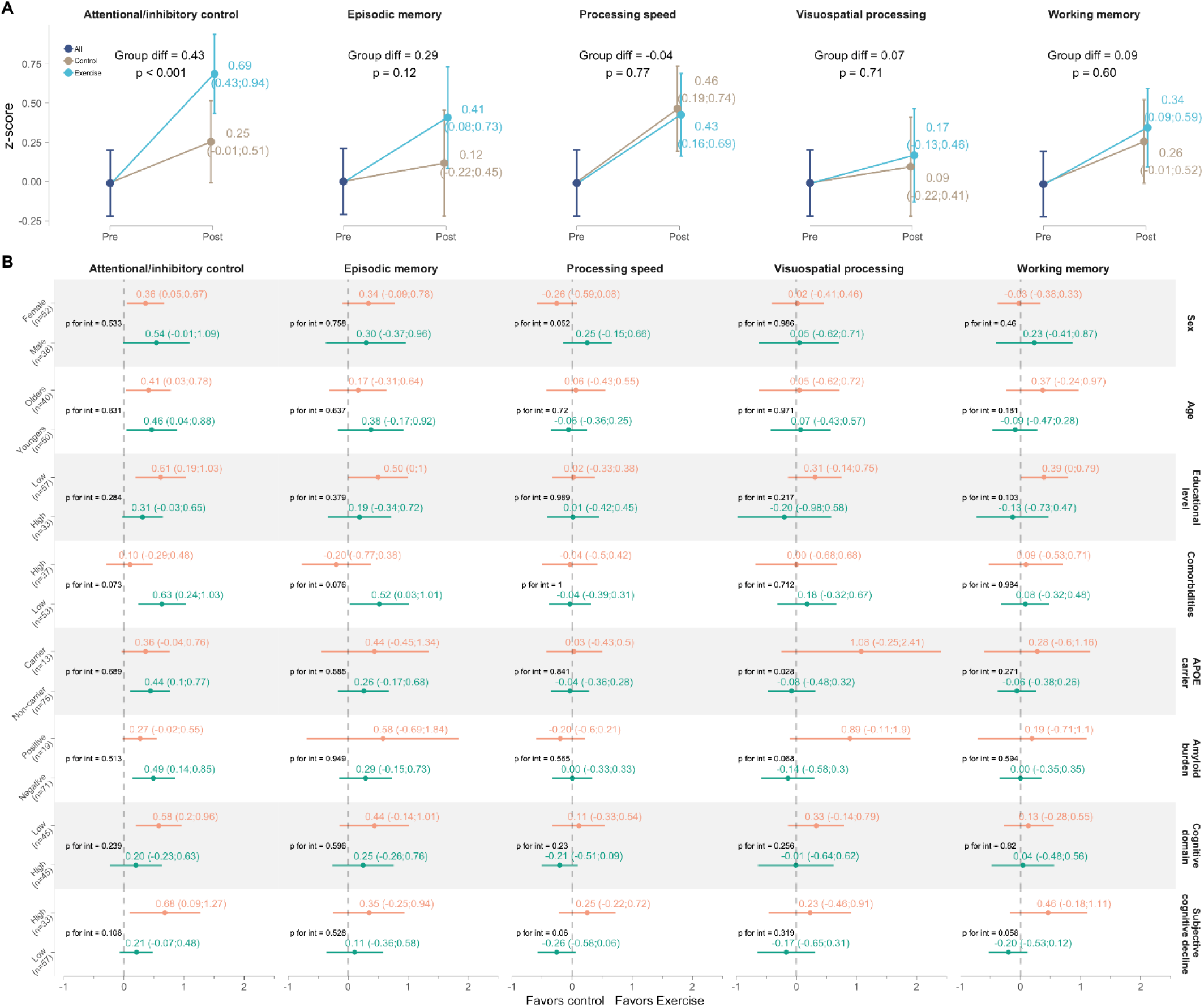
Intention-to-treat effects of the 24-weeks resistance exercise intervention on cognitive domains for the whole sample (A) and for subgroups (B). A. Dots indicate estimated marginal means at each time point (and 95% confidence intervals). B. Dots indicate mean difference of marginal means and 95% confidence intervals) between exercise and control in each subgroup.

### 3.4 Moderation analysis

Differences in RE effects on EF (**Figure 2C and Supplementary material 6, Table S11**) and other cognitive domains (**Figure 3B and Supplementary material 6, Table S12**) were examined as moderators at baseline (sex, age, educational level, comorbidities, APOE4 carriership, amyloid burden, baseline cognitive performance, and SCD). Greater effects on EF were found for participants reporting higher baseline levels of SCD (SMD = 0.47, 95% CI: - 0.01;0.94), in comparison with those reporting lower levels (SMD = −0.15, 95% CI: −0.53; 0.23; p for interaction = 0.043), for those with lower educational level (SMD = 0.41, 95% CI, 0.07; 0.75),in comparison with those reporting high educational level (SMD = −0.16, 95% CI, - 0.59;0.27; p for interaction = 0.05), and for the oldest participants (SMD = 0.45, 95% CI, 0.07;0.83), in comparation with the youngers (SMD = −0.01, 95% CI, −0.44;0.41; p for interaction = 0.11). Regarding the other cognitive domains, there were trends for moderation with various variables. Exercise consistently showed a trending moderation by baseline SCD in 3 out of the 5 cognitive domains: attentional/inhibitory control (p = 0.10), processing speed (p = 0.06), and working memory (p = 0.05).

### 3.5 Physical condition parameters

The RE effect on physical condition parameters is shown in **Figure 4A** (**Supplementary material 6, Table S13).** While the exercise group change more in all the parameters compared with control group, there were only significant differences between groups on the knee extension strength test (SMD = 0.25 z score, p = 0.02). No additional significant differences were detected (all p ≥ 0.05).

**Figure 4:**
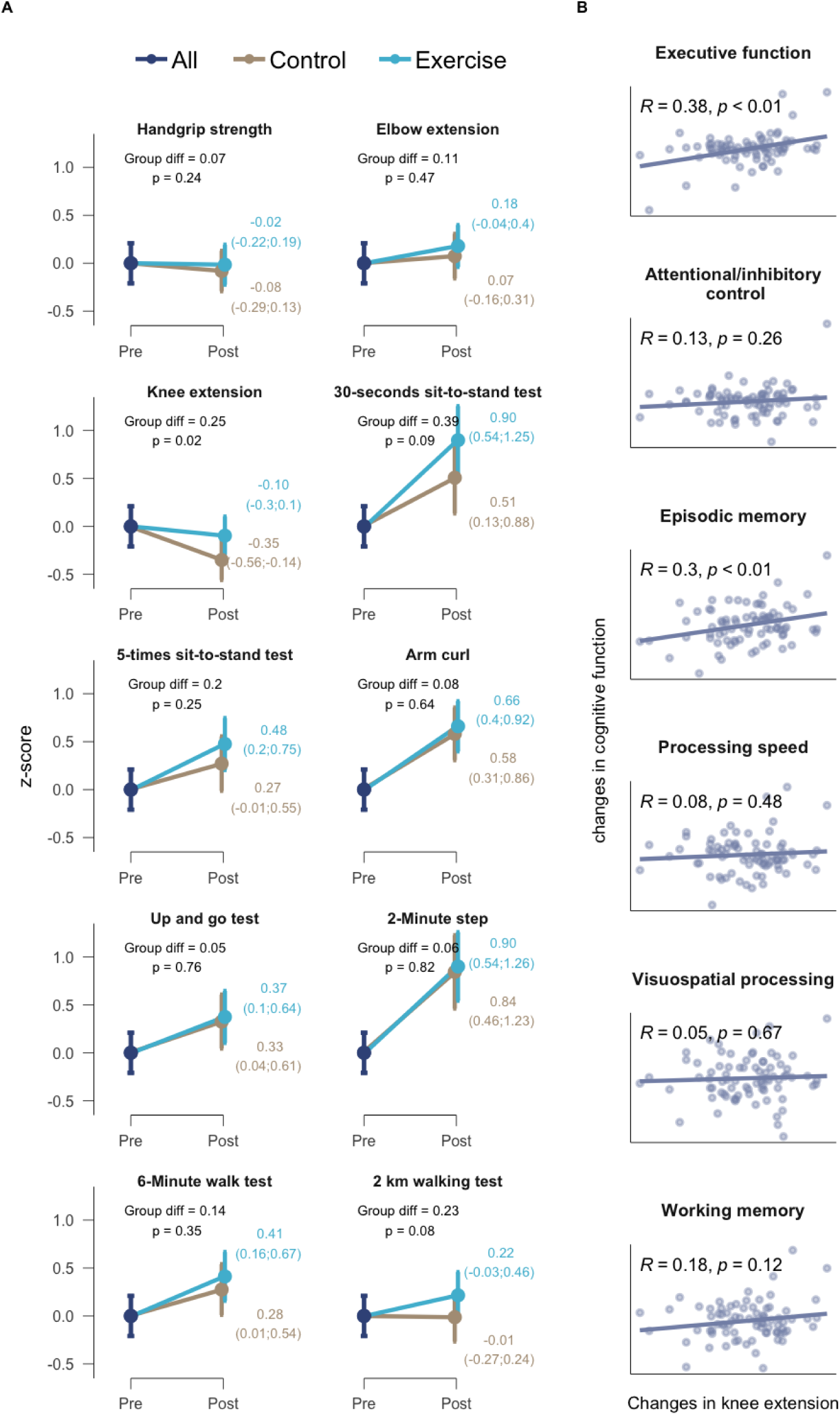
Intention-to-treat effects of the 24-weeks resistance exercise intervention on physical condition parameters (A) and correlation of Knee extension changes with cognitive function changes (B). A. Dots indicate estimated marginal means at each time point (and 95% Confidence Intervals). B. Dots indicate individual raw changes of each variable.

### 3.6 Post hoc mediation analysis and correlation between changes

Since we detected a significant effect of the exercise intervention on knee extension muscular strength test, we performed a mediation analysis to test whether changes to cognitive function were statistically mediated by changes in knee extension strength. No significant indirect effects were identified (**Supplementary material 6, Figure S3**). However, a positive correlations of changes in knee extension were found with changes in EF (R = 0.38, p < 0.001) and episodic memory (R = 0.31, p = 0.008) (**Figure 4B**), but not for the other cognitive domains (all p ≥ 0.12).

### 3.7 Intervention attendance and compliance

The average recorded attendance of the RE intervention group (46 participants) was 85.17%, including in-person (83.36%), and online sessions (1.81%). Moreover, 10% (53 out of 494 sessions) of the missed sessions were rescheduled. Two participants had less than 60% attendance, 4 participants between 60-80%, and 40 participants (87%) achieved >80% of attendance (**Figure 5A).** The mean RPE achieved was 5.3, with observed values ranging from 4.7 to 5.8 per session, and 4.6 (range: 3.8 to 5.4) per exercise (**Figure 5B**).

**Figure 5:**
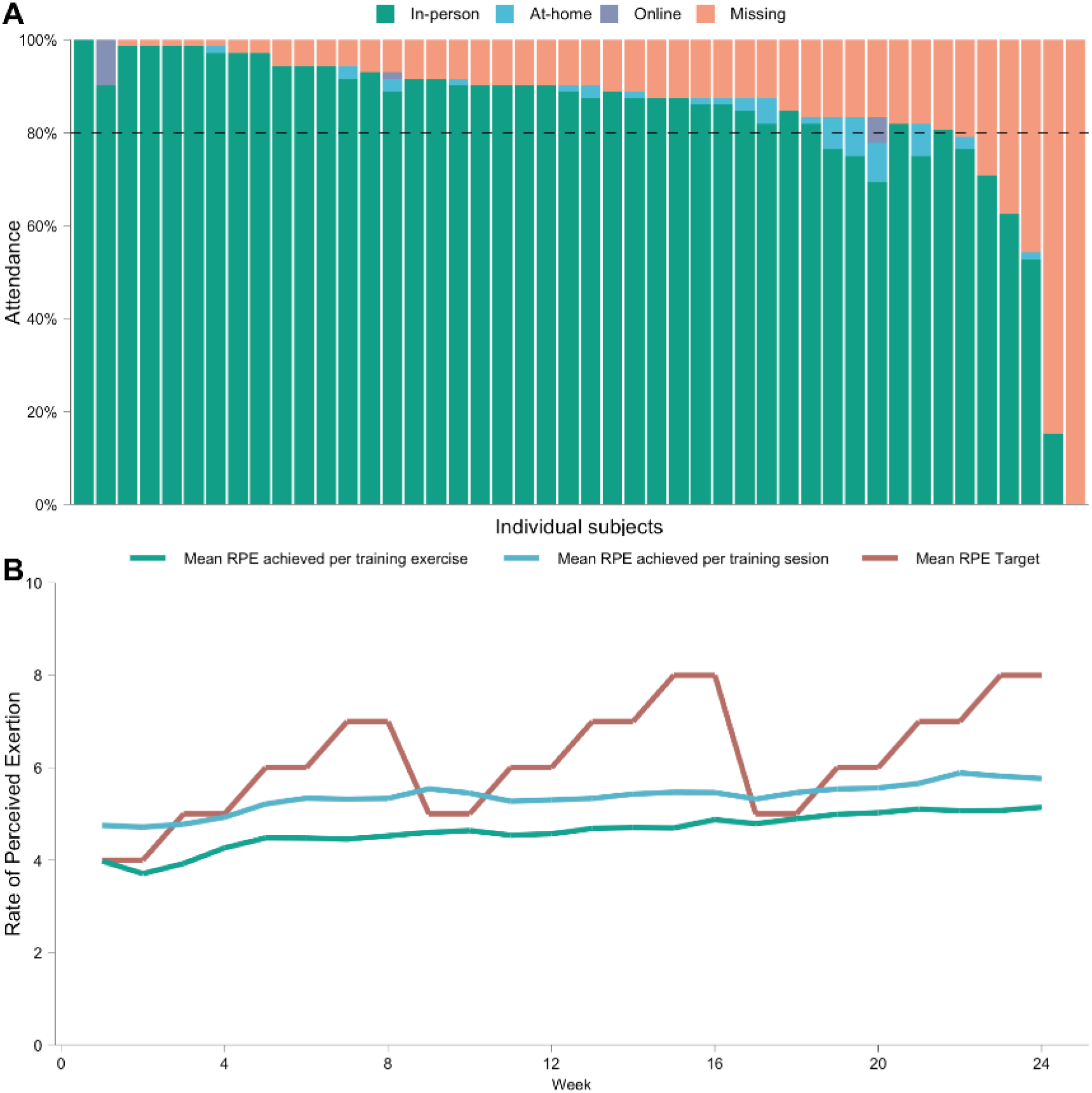
Attendance (A) and compliance (B) of the AGUEDA resistance exercise intervention.

### 3.8 Sensitivity analyses

Sensitivity analyses on our primary outcome including 12-week measurements showed that no differences were detected for EF at 24 weeks (SMD = 0.13, (95% CI, −0.16; 0.42), p = 0.37) (**Supplementary material 6, Figure S4**). The PP analysis on cognitive function (**Supplementary material 6, Figure S5**), which included only participants who attended more than 80% of the exercise intervention sessions (N = 85), showed similar results. This analysis compared the exercise group (those with >80% attendance) with the entire control group. RE group showed significantly better attention/inhibitory control than the control group at 24-weeks between groups (SMD = 0.41, p = 0.01). In addition, a sensitivity analyses with winsorized variables shows similar findings **(Supplementary material 6, Figure S6).**

### 3.9 Adverse event

A total of 8 adverse events were observed throughout the intervention period (**Supplementary material 6, Table S14**) with no causal relationship to the exercise intervention. In the RE group, 2 participants experienced humerus fractures, 1 had severe knee pain, and another showed reported intense back pain. Furthermore, one participant in this group underwent substantial fatigue, the cause of which eluded medical detection. On the other hand, the control group reported 2 adverse events, one participant had an ischemic stroke, while another was diagnosed with cancer. Only these two participants were excluded from the adverse event statistical analysis due to the potential impact of their conditions on the primary outcome which yielded similar results (data not shown).

## 4. Discussion

The AGUEDA trial aimed to assess the effects of a 24-week RE intervention on cognitive function in cognitively normal older adults. The ITT analysis revealed no significant differences in EF or other cognitive domains after 24 weeks. However, participants in the RE group demonstrated significantly greater improvements in attentional/inhibitory control compared to the control group. Subgroup analyses revealed a medium-sized but significant improvement in EF for those presenting high levels of SCD (i.e., memory complaints), and a small improvement for those with fewer years of education and the oldest adults. Significant gains in lower body muscular strength were observed and were positively correlated with changes in EF and episodic memory, although no significant statistical mediation effects were detected. These findings suggest that memory complaints, education, and age may moderate the cognitive effects of RE, underscoring the need for personalized interventions in at-risk groups.

The RE showed no significant group differences in most cognitive domains, but revealed selective attentional/inhibitory control improvements (i.e., Flanker Test, Stroop Test, DCCST, TMT) in both ITT and PP analyses. Previous systematic reviews have reported beneficial effects of RE on EF (28,82–87), attentional/inhibitory control (84,88), episodic memory (89,90), or working memory (91). Additionally, some RCTs specifically using elastic bands have reported cognitive benefits in general cognition but assessed through one or two single tests (23,92,93).

However, these systematic reviews emphasize important limitations that may affect the detection of REs cognitive benefits. One key limitation is the lack of clarity in identifying the specific neuropsychological tests and cognitive domains being evaluated (94). For instance, variations in the measurement of EF or other cognitive domains across different scales introduce greater variability and make interpretation of results more difficult (95). Furthermore, the role of neurobiological mechanisms unrelated to the specific cognitive demands of RE (28). Finally, the absence of standardized reporting on exercise intervention characteristics complicates accurate comparisons across RCTs, even among different types of RE interventions (i.e. elastic bands versus traditional weight training) (94).

To note, in line with our results, the most consistent benefits appear to be linked to inhibitory control (96,97), as demonstrated in prior interventions using aerobic-based exercise programs (98) or resistance-based exercise programs (99,100). The selective cognitive improvements of attention/inhibitory control could be attributed to the dual physical and cognitive demands of RE (101,102). Firstly, attentional/inhibitory control involved the dorsolateral prefrontal cortex (DLPFC) and anterior cingulate cortex (ACC), regions critical for decision-making and behavioral regulation (103); meanwhile RE may enhance activation and neuroplasticity in these areas, leading to more efficient neural processing (104,105). Secondly, RE may selectively enhance cognitive abilities that are most engaged during exercise (28) (e.g., continuous focus on movements, monitor body positioning, and adjust effort accordingly). These sustained attention demands not only improve the ability to concentrate on specific stimuli but also help individuals overcome physical discomfort and maintain effort, ultimately strengthening executive control mechanisms (106), particularly inhibitory control. Additionally, the overlap between motor learning and cognitive control is likely to contribute to these selective benefits (107). Emerging research shows that coordination training or cognitive-motor and dual-task training are particularly effective in promoting neuroplasticity (108–110), linked with RE, which involves motor tasks requiring planning, sequencing, and coordination of multiple body parts, skills that closely integrate with inhibitory control processes (111). Unlike cognitive functions such as working memory or processing speed, which often involve more passive cognitive processes (e.g., automatically following a familiar movement pattern like walking), RE requires the conscious inhibition of automatic reactions (e.g., maintaining proper posture or ensuring correct technique), a core aspect of inhibitory control (112). For example, managing fatigue, controlling intensity, or the balance and coordination RE with elastic bands engage inhibitory regulation, potentially leading to domain-specific cognitive gains (113). Neuroimaging evidence further supports these selective benefits; For example, the study by Liu-Ambrose et al. (2012) showed that 12 months of twice-weekly machine-based RE in older women resulted in improvements in inhibitory processes, which co-occurred with functional changes in hemodynamic activity in regions of the cortex commonly associated with response inhibition processing in flanker-type tasks (e.g., left anterior insula and the middle temporal gyrus) (114). In line with this, a previous systematic review (27) emphasized the importance of identifying cognitive domains that are most sensitive to exercise-induced improvements. This approach could facilitate the identification of specific individual or exercise characteristics which may be particularly responsive to exercise-mediated enhancements in cognitive function (27). These findings underscore the potential of RE as a targeted intervention for enhancing inhibitory control, highlighting the need for further research to optimize training protocols that maximize cognitive benefits.

Subgroup analyses revealed pronounced effects on the EF score in specific subgroups, such as individuals with lower education levels (≤12 years), oldest adults (≥72 years), and those reporting higher SCD (≥ 3 points of memory complaints). Indeed, the severity of self-reported memory complaints also moderates the effects of the AGUEDA RE intervention on 3 out of the 5 remaining cognitive domains (i.e., attentional/inhibitory control, processing speed, and working memory). These findings align with prior research showing greater responsiveness to exercise in high-risk populations for cognitive decline (e.g., oldest participants, ≥ 80 years (38); those with lower educational levels or those with higher SCD (115)). Furthermore, our results reinforce the idea that SCD itself may serve as a preclinical marker for identifying individuals who are particularly responsive to interventions targeting cognitive decline (85,116). Previous research has both supported and questioned the direct interaction between exercise and SCD-related cognitive changes (85) showing how early intervention in SCD could potentially improve the functioning and slow cognitive decline (117). Some speculative hypotheses, may explain why individuals with greater SCD benefit the most from exercise interventions: First, exercise enhances cognitive reserve and neural plasticity (118), increases brain volume (119), and improves neural efficiency, particularly benefiting those with higher SCD, whose cognitive reserve is more vulnerable and can be further influenced by modifiable factors such as education (120), Second, physical activity enhances social engagement, promotes emotional stability, and reduces distress (121), which is especially relevant for individuals with SCD, who often experience social withdrawal (122). Third, since individuals with higher SCD may have less efficient cognitive processing, exercise can help them allocate cognitive resources more effectively, leading to improved cognitive performance (123). This evidence highlights the potential of personalized exercise interventions tailored to individual characteristics, such as those with memory complaints, even before reaching a clinical phase, paving the way for tailored preventive strategies.

Our data suggests that the performance benefits of the AGUEDA RE are more pronounced in parameters involving lower-body strength, specifically in knee extension. Previous meta-analysis showed consistently positive effects between RE with elastic bands and muscle strength (124,125). Notably, handgrip strength has been identified as a potential marker for monitoring cognitive changes and predicting cognitive decline (126,127), however Zammit et al. (2018) emphasize the need for further investigation using multi-study approaches to evaluate the reliability and magnitude of this association (128). On the other hand, some studies on RE with elastic bands suggest that lower-limb exercises yield greater improvements in physical function parameters (23). This may be due to the accelerated decline in lower-body muscle strength with aging (129) and its established association with EF and general cognitive performance (130). Our findings further support this observation, revealing a positive correlation between improvements in lower-body strength and gains in EF and episodic memory, yet no evidence for mediation pathways was found. While these findings may underscore the potential role of physical function, particularly lower-body strength, as a determinant of cognitive health (131), larger elastic bands RE-based RCTs should confirm its mediation pathways during aging.

Exercise related-parameters —frequency, intensity, duration or type— are crucial components of intervention efficacy (13). In the AGUEDA intervention, the exercise group received a relatively high than usual attendance, as evidenced by an average rate of over 85% of the total prescribed sessions (61.2 out of 72 hours), comparing to previous reports with average of attendance rate of 70% (CI, 69–73%) (132,133). Additionally, over 87% of participants achieved the predefined PP attendance threshold of 80%. The average session intensity, measured by RPE, was 5.4 out of 10, aligning with the weekly target range of 4 to 8 based on the prescribed intensity. Both attendance and intensity may have played a role in the observed effects, as evidence suggests a connection between training volume and improvements in inhibitory control (134). In this context, Gomes et al. (2018) suggested that at least 52 hours of exercising (e.g., RE, aerobic exercise, mind-body or combinations) could improve cognitive performance, particularly in attentional and inhibitory control, in older adults (27). This may explain the lack of detectable EF improvements in the exercise group at 12 weeks, when only 36 hours of exercise were offered (with an average of ≈30 hours completed), but a greater tendency for gains compared to the control group observed at 24 weeks, with 72 hours offered (61.2 hours completed). These findings suggest that certain cognitive domains may require longer-duration interventions to achieve meaningful improvements (27,95).

This study had limitations, such as a small sample size for testing moderation or mediation, which may have led to being underpowered for these specific analyses. However, the sample size was adequate for the main analyses. Additionally, participant recruitment took place during the COVID-19 pandemic, which may have influenced the results (e.g., mask usage during some RE sessions and variability in participants’ prior experience and physical condition). It is important to note that one of the exclusion criteria was a prior COVID-19 diagnosis requiring hospitalization in an intensive care unit, and recruitment was conducted smoothly despite the pandemic context. Another limitation, as noted in previous systematic reviews, is the lack of a validated method for directly measuring and quantifying the external load when using elastic bands by an objectively assess (124); this is an issue we plan to explore in future research, based on our recently proposed load quantification equations (135). However, this study had several strengths, including the RCTs design and the use of elastic bands and body weight requiring minimal equipment and allowing for scalability. High adherence rates further underscore the program’s practicality. Additionally, comprehensive neuropsychological assessment, with multiple tests within each cognitive subdomain, and the exceptional phenotyping of the sample across all domains, are notable strengths of this study.

## 5. Conclusion

While no significant differences between group improvements were found in EF or other cognitive domains, this 24-week RE intervention led to selective cognitive benefits, particularly in attentional/inhibitory control. These findings align with existing evidence suggesting that the cognitive effects of RE may vary across different cognitive domains and be influenced by individual participant’s characteristics. This supports the value of personalized exercise interventions tailored to individual risk-populations, particularly in those with higher SCD.

In practice, this RE intervention offered a simple, safe, feasible, adaptable to real-world setting, low-cost and non-pharmaceutical approach to mitigate age-related cognitive decline (136). The program’s adaptability underscores its potential to contribute to public health strategies aimed at preserving cognitive function and promoting healthy aging, especially in high-risk populations. Future research should explore the dose-response relationship, extend intervention duration, and investigate the mechanisms underpinning these selective effects to optimize the design of a tailored exercise intervention for improving cognitive health.

## 6. Ethical Approval and consent to participate

Institutional review board approval was obtained from Andalusian Health Service prior to the start of the trial. CEIM/CEI Provincial de Granada; #2317-N-19; approval date: 25/05/2020.

## 7. Funding

This work was supported by grant RTI2018-095284-J-100 funded by MCIN/AEI/10.13039/501100011033/ and “ERDF Away of making Europe”, and RYC2019-027287-I funded by MCIN/AEI/10.13039/501100011033/ and “ESF Investing in your future”. P.S-U, M.O-R, A.C-P, I.M-F, J.F-O and L.S-A were supported by the Spanish Ministry of Science, Innovation and Universities (Margarita Salas, FPU 22/02476, FPU21/02594, JDC2022-049642-I, FPU22/03052 and FPU21/06192, respectively). AT was supported by the Junta de Andalucía, Spain, under the Postdoctoral Research Fellows (Ref. POSTDOC_21_00745). EAB was supported by the European Union’s Horizon 2020 research and innovation program under the Marie Skłodowska-Curie grant agreement No (101064851). JSM is supported by the National Agency for Research and Development (ANID)/Scholarship Program/DOCTORADO BECAS CHILE/2022–(Grant N°72220164). EAB has received funding from the European Union’s Horizon 2020 research and innovation programme under the Marie Skłodowska-Curie grant agreement No [101064851]. This work is part of Ph.D. Thesis conducted in the Biomedicine Doctoral Studies of the University of Granada. B-FG is supported by MCIN/AEI/10.13039/501100011033 and FSE+ (PID2022-137399OB-I00).

## 8. Conflict of Interest

The authors declare that they have no conflict of interest.

## 9. Patient and public involvement

The sponsors had no role in the design and conduct of the study, in the collection, analysis, and interpretation of data, in the preparation of the manuscript, or in the review or approval of the manuscript.

## Data Availability

All data produced in the present study are available upon reasonable request to the authors

https://github.com/aguedaprojectugr

## Acknowledgements

The authors would like to thank the participants who participated in this study for their time and interest.

---

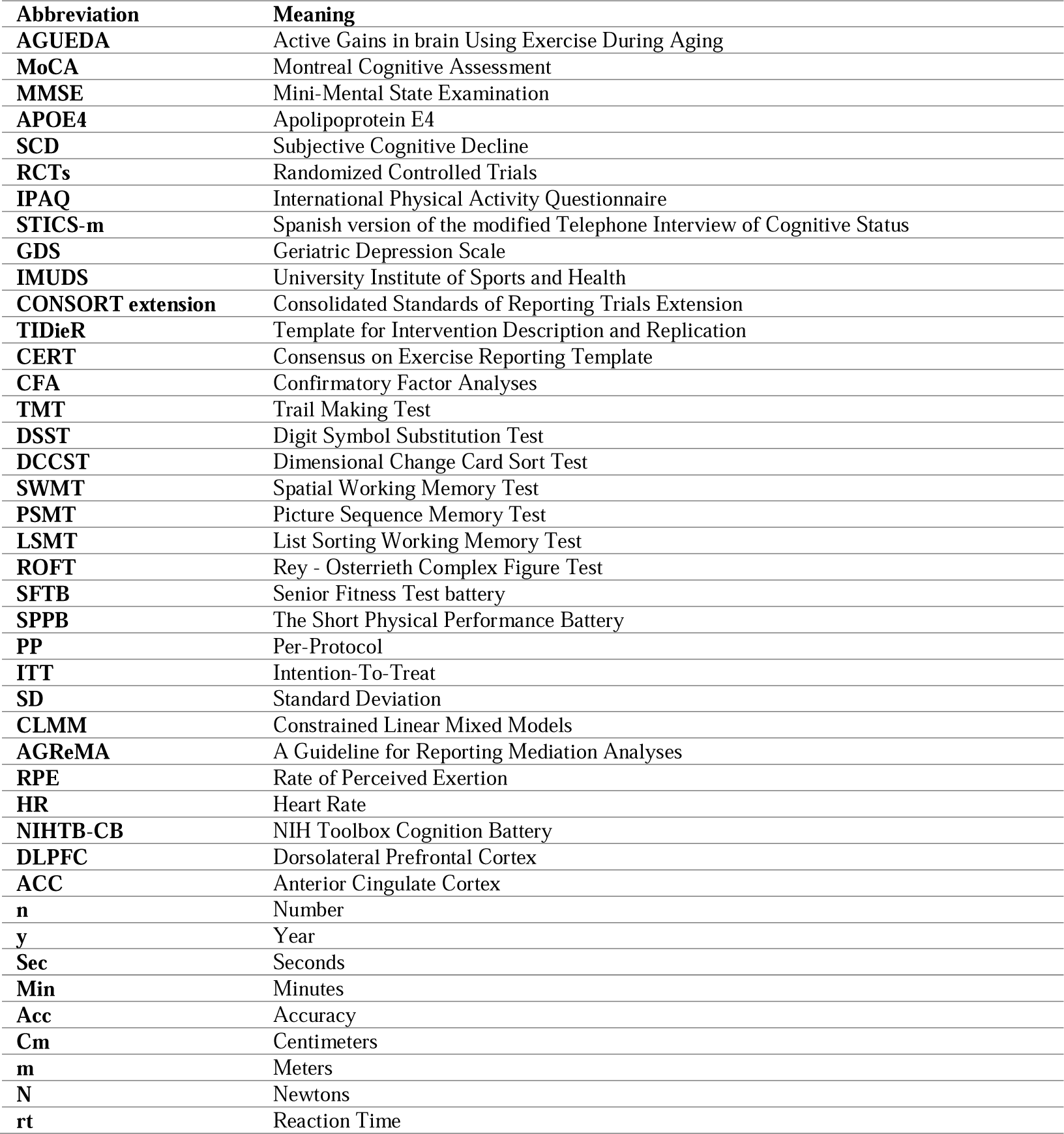

| **Abbreviation** | **Meaning** |
| --- | --- |
| **AGUEDA** | Active Gains in brain Using Exercise During Aging |
| **MoCA** | Montreal Cognitive Assessment |
| **MMSE** | Mini-Mental State Examination |
| **APOE4** | Apolipoprotein E4 |
| **SCD** | Subjective Cognitive Decline |
| **RCTs** | Randomized Controlled Trials |
| **IPAQ** | International Physical Activity Questionnaire |
| **STICS-m** | Spanish version of the modified Telephone Interview of Cognitive Status |
| **GDS** | Geriatric Depression Scale |
| **IMUDS** | University Institute of Sports and Health |
| **CONSORT extension** | Consolidated Standards of Reporting Trials Extension |
| **TIDieR** | Template for Intervention Description and Replication |
| **CERT** | Consensus on Exercise Reporting Template |
| **CFA** | Confirmatory Factor Analyses |
| **TMT** | Trail Making Test |
| **DSST** | Digit Symbol Substitution Test |
| **DCCST** | Dimensional Change Card Sort Test |
| **SWMT** | Spatial Working Memory Test |
| **PSMT** | Picture Sequence Memory Test |
| **LSMT** | List Sorting Working Memory Test |
| **ROFT** | Rey - Osterrieth Complex Figure Test |
| **SFTB** | Senior Fitness Test battery |
| **SPPB** | The Short Physical Performance Battery |
| **PP** | Per-Protocol |
| **ITT** | Intention-To-Treat |
| **SD** | Standard Deviation |
| **CLMM** | Constrained Linear Mixed Models |
| **AGReMA** | A Guideline for Reporting Mediation Analyses |
| **RPE** | Rate of Perceived Exertion |
| **HR** | Heart Rate |
| **NIHTB-CB** | NIH Toolbox Cognition Battery |
| **DLPFC** | Dorsolateral Prefrontal Cortex |
| **ACC** | Anterior Cingulate Cortex |
| **n** | Number |
| **y** | Year |
| **Sec** | Seconds |
| **Min** | Minutes |
| **Acc** | Accuracy |
| **Cm** | Centimeters |
| **m** | Meters |
| **N** | Newtons |
| **rt** | Reaction Time |

## Notes

### Competing Interest Statement

The authors have declared no competing interest.

### Clinical Trial

NCT05186090

### Clinical Protocols

https://pmc.ncbi.nlm.nih.gov/articles/PMC10239947/

https://pubmed.ncbi.nlm.nih.gov/37960912/

### Author Declarations

The trial protocol followed the principles of the Declaration of Helsinki and was approved by the Research Ethics Board of the Andalusian Health Service (CEIM/CEI Provincial de Granada; #2317-N-19) on May 25th, 2020

