## Supplemental files for "Effect of a 24-week resistance exercise intervention on cognitive function in cognitively normal older adults: The AGUEDA Randomized Controlled Trial"

**Supplemental material 1. CONSORT (Consolidated Standards of Reporting Trials) and TIDieR (Template for Intervention Description and Replication) checklists**

Table S1. CONSORT (Consolidated Standards of Reporting Trials) checklist

| **Section/Topic** | **Item No** | **Checklist item** | **Reported on Page No** |
| --- | --- | --- | --- |
| **Title and abstract** | | |  |
|  | 1a | Identification as a randomized trial in the title | 0 |
|  | 1b | Structured summary of trial design, methods, results, and conclusions (for specific guidance see CONSORT for abstracts) | 0 |
| **Open science** | | | |
| Trail registration | 2 | Name of trial registry, identifying number (with URL) and date of registration | 3 |
| Protocol and statistical analysis plan | 3 | Where the trial protocol and statistical analysis plan can be accessed | 3 |
| Data sharing | 4 | Where and how the individual de-identified participant data (including data dictionary), statistical code and any other materials can be accessed | 3 |
| Funding and conflicts of interest | 5a | Sources of funding and other support (eg, supply of drugs), and role of funders in the design, conduct, analysis and reporting of the trial | 20 |
|  | 5b | Financial and other conflicts of interest of the manuscript authors | 21 |
| **Introduction** | | | |
| Background and rationale | 6 | Scientific background and rationale | 1 |
| Objectives | 7 | Specific objectives related to benefits and harms | 2 |
| **Methods** | | | |
| Patient and public involvement | 8 | Details of patient or public involvement in the design, conduct and reporting of the trial | 3 |
| Trial design | 9 | Description of trial design including type of trial (eg, parallel group or crossover), allocation ratio, and framework (for example, superiority, equivalence, non-inferiority or exploratory) | 3, Fig. 1 |
| Changes to trial protocol | 10 | Important changes to the trial after it commenced including any outcomes or analyses that were not prespecified, with reason | 10, Fig.1 |
| Trial setting | 11 | Settings (such as community or hospital) and locations (eg, countries or sites) where the trial was conducte | 3 |
| Eligibility criteria | 12a | Eligibility criteria for participants | 4, Suppl.2(Table S3) |
|  | 12b | If applicable, eligibility criteria for sites and for individuals delivering the interventions (eg, surgeons or physiotherapists) | NA |
| Intervention and comparator | 13 | Intervention and comparator with sufficient details to allow replication. If relevant, where additional materials describing the intervention and comparator (eg, intervention manual) can be accessed | 3 and Ref.49 |
| Outcomes | 14 | Prespecified primary and secondary outcomes, including the specific measurement variable (eg, systolic blood pressure), analysis metric (for example, change from baseline, final value, time to event), method of aggregation (eg, median, proportion), and time point for each outcome | 5,6 Suppl. 3, 4 and 5 |
| Harms | 15 | How harms were defined and assessed (eg, systematically or non-systematically) | NA |
| Sample size | 16a | How sample size was determined, including all assumptions supporting the sample size calculation | Ref. 43 |
|  | 16b | Explanation of any interim analyses and stopping guidelines | NA |
| Randomization | | | |
| Sequence generation | 17a | Who generated the random allocation sequence and the method used | Ref. 43 |
|  | 17b | Type of randomization and details of any restriction (eg, stratification, blocking and block size) | Ref. 43 |
| Allocation concealment mechanism | 18 | Mechanism used to implement the random allocation sequence (eg, central computer/telephone; sequentially numbered, opaque, sealed containers), describing any steps to conceal the sequence until interventions were assigned | Ref. 43 |
| Implementation | 19 | Whether the personnel who enrolled and those who assigned participants to the interventions had access to the random allocation sequence | Ref. 43 |
| Blinding | 20a | Who was blinded after assignment to interventions (eg, participants, care providers, outcome assessors, data analysts) | Ref. 43 |
|  | 20b | If blinded, how blinding was achieved and description of the similarity of interventions | Ref. 43 |
| Statistical methods | 21a | Statistical methods used to compare groups for primary and secondary outcomes, including harms | 8 |
|  | 21b | Definition of who is included in each analysis (eg, all randomized participants), and in which group | 8 |
|  | 21c | How missing data were handled in the analysis | 8 |
|  | 21d | Methods for any additional analyses (eg. subgroup and sensitivity analyses), distinguishing prespecified from post hoc | 8 |
| **Results** | | | |
| Participant flow, including flow diagram | 22a | For each group, the numbers of participants who were randomly assigned, received intended intervention, and were analyzed for the primary outcome | 3, Fig. 1 |
|  | 22b | For each group, losses and exclusions after randomization, together with reasons | Fig. 1 |
| Recruitment | 23a | Dates defining the periods of recruitment and follow-up for outcomes of benefits and harms | 3 |
|  | 23b | If relevant, why the trial ended or was stopped | NA |
| Intervention and comparator delivery | 24a | Intervention and comparator as they were actually administered (eg, where appropriate, who delivered the intervention/ comparator, how participants adhered, whether they were delivered as intended (fidelity)) | 12 |
|  | 24b | Concomitant care received during the trial for each group | 13 |
| Baseline data | 25 | A table showing baseline demographic and clinical characteristics for each group | 3, Fig.1, Suppl.6(Table S10) |
| Numbers analyzed, outcomes and estimation | 26 | For each primary and secondary outcome, by group:  • the number of participants included in the analysis  • the number of participants with available data at the outcome time point  • result for each group, and the estimated effect size and its precision (such as 95% confidence interval)  • for binary outcomes, presentation of both absolute and relative effect size | 10, Fig.1, Fig.2A, Fig.2B, Fig.3A,  Suppl.6(Table S13, Fig.S3, Fig.S4, Fig.S5, Fig. S6) |
| Harms | 27 | All harms or unintended events in each group | NA |
| Ancillary analyses | 28 | Any other analyses performed, including subgroup and sensitivity analyses, distinguishing pre-specified from post hoc | 10, Fig.2C, Fig.3B, Fig.4,  Suppl.6(S11, S12) |
| **Discussion** | | | |
| Interpretation | 29 | Interpretation consistent with results, balancing benefits and harms, and considering other relevant evidence | 14 |
| Limitations | 30 | Trial limitations, addressing sources of potential bias, imprecision, generalisability, and, if relevant, multiplicity of analyses | 19 |

Table S2. TIDieR (Template for Intervention Description and Replication) checklist

| **Item number** | **Item** | **Where located** | |
| --- | --- | --- | --- |
|  |  | **Primary Paper (page)** | **Supplementary material** |
| **1.** | **BRIEF NAME** | 0 |  |
|  | Provide the name or a phrase that describes the intervention. |  |  |
| **2.** | **WHY** | 1 |  |
|  | Describe any rationale, theory, or goal of the elements essential to the intervention. |  |  |
| **3.** | **WHAT**  Materials: Describe any physical or informational materials used in the intervention, including those provided to participants or used in intervention delivery or in training of intervention providers. Provide information on where the materials can be accessed (e.g. online appendix, URL). | 4,8 | Ref.49 |
| **4.** | **PROCEDURES** | 4,8 | Ref.49 |
|  | Describe each of the procedures, activities, and/or processes used in the intervention, including any enabling or support activities. |  |  |
| **5.** | **WHO PROVIDED** | 4 | Ref.49 |
|  | For each category of intervention provider (e.g. psychologist, nursing assistant), describe their expertise, background and any specific training given. |  |  |
| **6.** | **HOW** | 4 | Ref.49 |
|  | Describe the modes of delivery (e.g. face-to-face or by some other mechanism, such as internet or telephone) of the intervention and whether it was provided individually or in a group. |  |  |
| **7.** | **WHERE** | 4 | Ref.49 |
|  | Describe the type(s) of location(s) where the intervention occurred, including any necessary infrastructure or relevant features. |  |  |
| **8.** | **WHEN and HOW MUCH** | 4 | Ref.49 |
|  | Describe the number of times the intervention was delivered and over what period of time including the number of sessions, their schedule, and their duration, intensity or dose. |  |  |
| **9.** | **TAILORING** | 4 | Ref.49 |
|  | If the intervention was planned to be personalized, titrated or adapted, then describe what, why, when, and how. |  |  |
| **10.** | **MODIFICATIONS** | 4 | Ref.49 |
|  | If the intervention was modified during the course of the study, describe the changes (what, why, when, and how). |  |  |
| **11.** | **HOW WELL** | 13 | Ref.49 |
|  | Planned: If intervention adherence or fidelity was assessed, describe how and by whom, and if any strategies were used to maintain or improve fidelity, describe them. |  |  |
| **12.** | Actual: If intervention adherence or fidelity was assessed, describe the extent to which the intervention was delivered as planned. | 13 | Ref.49 |

**Supplemental material 2. AGUEDA inclusion and exclusion criteria for selecting participants**

| Table S3. AGUEDA inclusion and exclusion criteria for selecting participants. | |
| --- | --- |
| **Inclusion criteria** | **Exclusion criteria** |
| - Men and women 65-80 years old. | - Ambulatory with pain or regular use of an assisted walking device. |
| - Able to speak and read fluent Spanish. | - Medical contraindication for inclusion in an exercise program. |
| - Living in community settings during the study. | - Neurological condition (Multiple Sclerosis, Parkinson Disease, Dementia) or brain injury (traumatic or stroke). |
| - Reliable means of transportation. | - Current diagnosis and treatment of a DSM V Axis I or II disorder including major depression, and seeing a psychologist, therapist, or psychiatrist in the last year. |
| - Being physically inactive: (i) not participating in any resistance exercise programs in the last 6 months, and (ii) accumulating less than 600 METs/week of moderate-vigorous physical activity. | - History of major psychiatric illness including schizophrenia, general anxiety disorder or depression (GDS-30≥15). |
| - Classified as cognitively healthy according to STICS-m, MoCA and MMSE. | - Current treatment for congestive heart failure, angina, uncontrolled arrhythmia, deep venous thrombosis or another cardiovascular event. |
|  | - Myocardial infarction, coronary artery bypass grafting, angioplasty or other cardiac condition in the last year. |
|  | - Current or previous treatment for any type of cancer. |
|  | - Type I Diabetes or uncontrolled Type II Diabetes defined as insulin dependent. |
|  | - Current treatment for alcohol or substance abuse. |
|  | - Presence of metal implants (e.g., pacemaker, stents, joint replacement) that would be MRI ineligible. |
|  | - Claustrophobia. |
|  | - Color blindness. |
|  | - Diagnosis of COVID-19 with hospitalization in intensive care unit. |
|  | - Any other consideration that interferes with the study aims and could be a risk to the participant, at the discretion of the researcher |
| DSM: Diagnostic and Statistical Manual of Mental Disorders, GDS: Geriatric Depression Scale, METs: Metabolic Equivalents, MMSE: Mini-Mental State Examination; MoCA: Montreal Cognitive Assessment. STICS-m: Modified Spanish Telephone Interview of Cognitive Status; MRI: Magnetic resonance imaging. COVID-19: Coronavirus disease-19 | |

**Supplementary material 3. Cognitive outcomes**

**1. Description of the cognitive test**

Participants completed a comprehensive neuropsychological evaluation using both paper-based and computer-based assessments. All computerized cognitive tests were administered via PC or iPAD with a standard monitor and keyboard. Each test is described below.

***1.1 Montreal Cognitive Assessment***

The Montreal Cognitive Assessment (MoCA) is a widely used and validated screening instrument for detecting cognitive impairment (60). The MoCA involves brief assessments of short-term memory, visuospatial abilities, orientation, attention, language, and executive functions, each of which is scored separately. These scores can be summed to calculate a total score or used as individual scores for each section. Participants can receive up to 30 points on the MoCA. The Clock Drawing Test is part of the MoCA and assesses visuospatial and planning abilities. Participants were instructed to draw a clock, including placing the hands to represent a specific time (e.g., 11:10). Additionally, the MoCa delayed recall was measured to assess participants ability to remember a list of words after a short delay. If they couldn’t recall the words, multiple-choice cues were provided to help them remember (maximum score 3 points).

***1.2 Wechsler Adult Intelligence Scale***

The Wechsler Adult Intelligence Scale (WAIS) is a paper-based test that provides a detailed evaluation of intellectual abilities by assessing skills such as reasoning, problem-solving, memory, and processing speed (137). Two subtests from the WAIS were used:

(i) The Matrix Reasoning subtest from the WAIS evaluates visuospatial reasoning and problem-solving skills. Participants were shown a series of incomplete geometric patterns and were asked to select the option that completes the logical sequence, (ii) The Block Design subtest measures visuospatial processing and abstract reasoning. Participants were shown a series of visual patterns and asked to replicate them using colored cubes within a time limit. Accuracy and response times were recorded in both tests (e.g., higher scores reflect better cognitive performance)

***1.3 Trail Making Test***

The Trail Making Test (TMT) is a paper-and-pencil test that measures cognitive flexibility and set-shifting (54). TMT, Part A, is a measure of psychomotor processing speed that requires participants to connect numbers displayed on a page in ascending order. TMT, Part B, measures set-shifting and requires participants to alternate between numbers and letters to connect them in ascending order. Time to completion (seconds) was measured, and reverted (multiplied by -1) with higher values reflecting better performance.

***1.4 Digit Symbol Substitution Test***

The Digit Symbol Substitution Test (DSST) is a paper-and-pencil test designed to assess psychomotor processing speed and basic attention (55). The DSST requires copying as many novel symbols corresponding to numbers as possible in 120 seconds. Total number of correct responses was measured. Higher scores reflect better cognitive performance.

***1.5 Rey Auditory Verbal Learning Test***

The Rey Auditory Verbal Learning Test (RAVLT) assesses verbal episodic memory (61). Participants are presented with a list of words and are asked to recall them. The total learning correct score is the sum of the 5 consecutive learning trials from list A. The delayed recall score measures the number of words correctly recalled from List A after a 20-minute delay. The recognition score reflects the number of correct responses when participants identify words from the original list among new words. A recognition index was measured by the number of correct responses of the recognition trial minus the number of intrusions false correct responses of the recognition trial. Higher scores reflect better cognitive performance.

***1.6 Rey - Osterrieth Complex Figure Test***

The Rey-Osterrieth Complex Figure Test (ROFT) evaluates visual episodic memory and visuoconstructive skills (138). Participants are first asked to copy a complex geometric figure. Later, after a 3 minute delay, they are asked to reproduce the figure from memory. The number of correct elements recalled in both trials was recorded (maximum score of 36 points). Higher scores reflect better cognitive performance.

***1.7 Picture Sequence Memory Test***

Picture Sequence Memory Test (PSMT) is a computer-based test from NIH Toolbox Cognition Battery (version 1.17)(59), that evaluates episodic memory. Participants were presented with a series of pictures in a specific order, and their task was to remember the order in which the pictures were shown. The number of pictures the participants were required to order ranged from 15-picture sequences (First trial) to 18-picture sequences (Second trial). Raw scores reflect the cumulative number of adjacent pairs of pictures remembered correctly over the two trials. Higher scores reflect better cognitive performance.

***1.8 Dimensional Change Card Sort Test***

Dimensional Change Card Sort Test (DCCST) is a computer-based test from NIH Toolbox Cognition Battery (version 1.17) which assesses cognitive flexibility and the ability to shift attention between different dimensions or rules (59). Participants were trained on one rule and then required to shift their attention and adapt to a new sorting rule during the shift phase. In test trials, participants sorted cards based on the current rule presented on the screen, which could switch between shape and color. Accuracy and response times were recorded (e.g., higher scores reflect better cognitive performance)

***1.9 List Sorting Working Memory Test***

List Sorting Working Memory test (LSWMT) is a computer-based test from NIH Toolbox Cognition Battery (version 1.17) that assesses working memory (65). Participants were asked to recall and sequence different stimuli by size that were presented both visually and via audio across two trials. In 1-list trials, all stimuli were from the same category (foods or animals). In 2-list trials, the stimuli were from two different categories (foods and animals). The participant was asked to sequence the food stimuli followed by the animal stimuli. The total number of correct items across the 1-list and 2-list conditions (maximum score of 28) were recorded.

***1.10 Flanker Test***

The Flanker Test is an arrow version of the computer-based test from NIH Toolbox Cognition Battery (version 1.17) that measures inhibition (59). During this test, participants are presented with a series of five arrows displayed in a row on the screen, each pointing either left or right. Participants must respond as quickly as possible to the direction of the middle arrow (left or right) using the corresponding arrow keys on the keyboard, while ignoring the external arrows.

Accuracy and response times were recorded. Higher scores reflect better cognitive performance.

***1.11 Stroop Test***

The Stroop Test is a computerized measure of inhibitory control from E-prime v2.0 (Psychological Software Tools, Pittsburgh, PA, USA). Words were presented on the screen one at a time. The words are colour words ‘green’, ‘blue’, ‘red’, ‘yellow’ that are printed in these different coloured inks. The participant is required to press a key with the colour indicated on for the ink colour, inhibiting the written word presented. There were 36 practice trials (22 congruent trials; written word and ink colour the same, 14 incongruent trials ;written word and ink colour different) and 72 experimental trials (36 congruent, 36 incongruent). Accuracy and reaction times were recorded. Higher scores reflect better cognitive performance.

***1.12 Spatial Working Memory Test***

Spatial Working Memory Test (SWMT) is a computer-based test that assesses an individual's ability to temporarily store and manipulate spatial information from E-prime v2.0 (Psychological Software Tools, Pittsburgh, PA, USA) (139). The participants were asked to stare at a crosshair in the middle of the screen. Afterwards, 2, 3 or 4 black dots appear at random locations on the screen and disappear. Then, a red dot appears on the screen and the participant must respond if the dot either is or not placed in the same place as one of the black dots that appeared before as fast as possible. Accuracy and reaction times were recorded. Higher scores reflect better cognitive performance.

***1.13 Task Switching Test***

The Task-Switching Test computer test assessed the ability to flexibly switch the focus of attention between multiple task sets from E-prime v2.0 (Psychological Software Tools, Pittsburgh, PA, USA). Participants had to switch between judging whether a number was odd or even and judging whether it was higher or lower than 5. The eligible numbers were 1, 2, 3, 4, 6, 7, 8, and 9, and were presented individually for 1500 ms in the center of the computer screen, enclosed within either a circle or a square, with the constraint that the same number did not appear twice in succession. The shape of the frame determined the task rule: if the number was inside a circle, participants had to decide whether it was higher or lower than 5, whereas if the number was inside a square, they had to determine whether it was odd or even. Participants responded using their left hand for the high/low task (‘x’ key for high, ‘z’ key for low) and their right hand for the odd/even task (‘n’ key for odd, ‘m’ key for even). The test consisted of three blocks: a first block (24 trials) where all numbers appeared inside circles, requiring only the high/low classification; a second block (24 trials) where all numbers appeared inside squares, requiring only the odd/even classification; and a third mixed-task block (120 trials) where numbers appeared inside either circles or squares, requiring participants to switch between tasks based on the shape of the frame. In the switching block, trials were categorized as repeat trials (where the task remained the same as in the previous trial) or switch trials (where the task changed from the previous trial). The switch cost was calculated as the difference in reaction times between switch and repeat trials, reflecting the cognitive effort required to adapt to a new task rule. Reaction times, accuracy rates, and switch costs were recorded to assess cognitive flexibility and task-switching efficiency.

***1.14 N-back***

The N-back computer test is a measure of visual working memory from E-prime v2.0 (Psychological Software Tools, Pittsburgh, PA, USA) (66), which was completed by participants during their MRI scan. On the 1-back condition, participants were instructed to indicate whether the letter on the screen matched the letter that was previously displayed. The 2-back condition requires a greater cognitive load and instructed participants to indicate whether the letter on the screen matched the letter that was displayed two trials previously. Accuracy and reaction time for both conditions were recorded.

**2. Executive function score**

***2.1 Creation of composite executive function score***

*2.1.1 Methods*

Z-standardization was used to ensure that outcomes with different response scales were on a standardized and comparable metric. Inverse variables were used when needed so a higher value indicated better performance. Firstly, a preliminary data checking procedure was conducted: Means, minimums and maximums of raw grounded outcomes (i.e., reaction times and accuracies) of each cognitive indicator were inspected. Pearson correlations were performed to test how the cognitive indicators related to each other.

Secondly, analyses were conducted to ensure that assumptions of normality and linearity. Bartlett’s Sphericity Test assessed the probability that at least some of the variables were significantly correlated and the Kaiser-Meyer-Olkin (KMO) statistic assessed the factorability of the data using the R package performance (140). The KMO statistic, ranging from 0 to 1, predicted whether the data were likely to factor well given the correlations among the variables. Using Kaiser’s guidelines (141) a cutoff of KMO ≥ .60 was used.

The CFA was conducted using the “*lavaan”* package in R (142) with maximum likelihood estimation. We tested the fit of several models that differed in the following ways for creating the executive function score: i) whether model fit improved with three cognitive domains factors, ii) whether model fit improved with a second-order general cognitive factor and iii) whether the model fit was better with a first-order factor of executive function

Firstly, guided on available evidence and theoretical positions of executive function, a model that hypothesized three factors was tested. Specifically, a model with three cognitive domains factors: 1) Working memory derived from SWMT, PSMT and LSMT, 2) Inhibitory control from Flanker test and Stroop test and 3) Cognitive flexibility from Task switching, DCCST, DSST and TMT. Secondly, because of a bad fit of the previous model we performed a two-factor model and a second-order one dropping indicators with factor loadings lower than 0.45, specifically, with 1) working memory derived from SWMT, PSMT and LSMT and 2) Cognitive flexibility from DCCST, DSST, TMT and Flanker Test.

Finally, the first-order CFA with a general executive function factor was the one that best fit, which added task-specific covariance that allowed the scores within a task to correlate with one another, with loadings freely estimated (143). The latent scale was identified by fixing the loading of a reference indicator to one. The remaining pattern coefficients, factor variances, and factor covariances were freely estimated (144). Each criterion was unidimensional and loaded on only one factor. See Table S4 for a list of measures and outcomes analyzed.

Five different goodness-of-fit statistics were used to assess model fit using established cut-offs, including the χ2 (p-value ≥ .05) (145), the comparative fit index (CFI) (> .95), the root mean square error of approximation (RMSEA) (< .08) (146), the Standardized Root Mean Square Residual (SRMR) (< .08) (147,148), and indicators with factor loadings less than 0.45 were removed to improve model fit, as it has been suggested that a cut-off of 0.45 indicates good (30%) overlapping variance (149).

Table S4. Executive function Measures and conditions included in the hypothesized and final factor analysis models

| Measure | Outcome | Hypothesized | Final |
| --- | --- | --- | --- |
| Trail Making test (TMT) | Interference score  ( Time part B – Time part A) (sec) | ✔️ | ✔️ |
| Digit Symbol Substitution test (DSST) | Total correct score (n) | ✔️ | ✔️ |
| Dimensional Change Card Sort Test (DCCST) | Inverse efficiency score (switch trials - high load) (rt/acc) | ✔️ | ✔️ |
| Spatial Working Memory Test (SWMT) | Inverse efficiency score of all trials (rt/acc) | ✔️ | ✔️ |
| Picture Sequence Memory Test (PSMT) | Theta Score* | ✔️ |  |
| List Sorting Working Memory Test (LSWMT) | Total correct score (n) | ✔️ |  |
| Flanker Test | Inverse efficiency score of Incongruent trials (rt/acc) | ✔️ |  |
| Stroop Test | Inverse efficiency score of Incongruent trials (rt/acc) | ✔️ |  |
| Task Switching Test | Global cost score  (switch trials - non-switch trials) | ✔️ |  |

Abbreviations: acc, accuracy; n, number; sec, seconds; rt, reaction time. *Theta score is a statistical measure that reflects a participant's relative performance, similar to a z-score, which is a measure that indicates how far a participant's score is from the average (mean) of a normative group, in terms of standard deviations.

*2.1.2 Results*

*2.1.2.1 Data preprocessing and winsorization*

The 90 participants randomized were reviewed for consistency by checking the minimum and maximum values of the raw outcomes and evaluating the biological plausibility of the data to identify any implausible values. For sensitivity analyses, 27 outliers, defined as values exceeding four standard deviation from the mean, were winsorized by replacing them with the nearest non-extreme value (See Table S5). However, winsorization was not applied in the main analyses.

Table S5. Cognitive measure, outcome and the number of participants winsorized

| Cognitive measure | Outcome (unit) | Number of participants winsorized |
| --- | --- | --- |
| Rey Auditory Verbal Learning Test (RAVLT) | Recognition - Number of correct responses of the recognition trial (n) | 2 |
| Rey - Osterrieth Complex Figure Test (ROFT) | Raw score copy (n) | 1 |
| Trail Making Test (TMT) | Time part A (sec) | 3 |
|  | Time part B (sec) | 5 |
|  | Inverse efficiency score of all trials (rt/acc) | 5 |
| Dimensional Change Card Sort Test (DCCST) | Computed score* (n) | 4 |
| Dimensional Change Card Sort Test (DCCST) | Inverse efficiency score (switch trials - high load) (rt/acc) | 3 |
| Spatial Working Memory Test (SWMT) | Inverse efficiency score of all trials (rt/acc) | 4 |

Abbreviations: acc, accuracy; n, number; sec, seconds; rt, reaction time. *Theta score is a statistical measure that reflects a participant's relative performance, similar to a z-score, which is a measure that indicates how far a participant's score is from the average (mean) of a normative group, in terms of standard deviations. Winsorization was applied only in the sensitivity analyses and not in the main analyses.

*2.1.2.2 Missing data*

Of the 90 randomized participants, 87 successfully completed all the baseline cognitive assessments included in the executive function scores and other domains. 69 successfully completed all the cognitive assessments at both baseline and at 24-weeks. The number of missing data points from the original randomized sample at 24-weeks were as follows: for composite executive function score, attentional/inhibitory control, episodic memory, and processing speed, the RE had n = 3 missing cases, while the control group had n = 6; for visuospatial processing and working memory, the RE had n = 3, and the control group had n = 7. A chi-square of the completion ratio between groups was not significant (all p > 0.43), suggesting the proportion of missing data did not differ by group assignment. Moreover, a comparison of the subsample who did not successfully complete the 24-week cognitive assessment to the sample who completed both baseline and 24-week cognitive assessments revealed no differences in sex and education (all p > 0.05). However, age was significantly related to missing data (p = 0.019), indicating that older participants were more likely to have missing follow-up cognitive test data. Based on these associations, our primary regression models were adjusted for the effect of age, assuming that missing data were missing at random (MAR), which includes covariate-dependent missing completely at random (MCAR) as a special case.

Table S6. Cognitive measure, missing data and chi-square

|  | **Missing data** | | **All chi-square** | **Sub-group chi-square** | | |
| --- | --- | --- | --- | --- | --- | --- |
|  | Exercise | Control | p-value | Sex | Age | Education |
| Composite executive function score and other domains | 3 | 6 | 0.439 | 0.656 | 0.019 | 0.840 |

Analysis of Chi-square were done with the lower n value.

*2.1.2.3 Pearson correlations between executive function measures.*

Correlations between the executive function indicators are presented in **Supplementary material 3, Figure S1**. All other scores were significantly correlated with each other (ranging between r = 0.15, and r = 0.55) but the task switching task was negatively correlated.

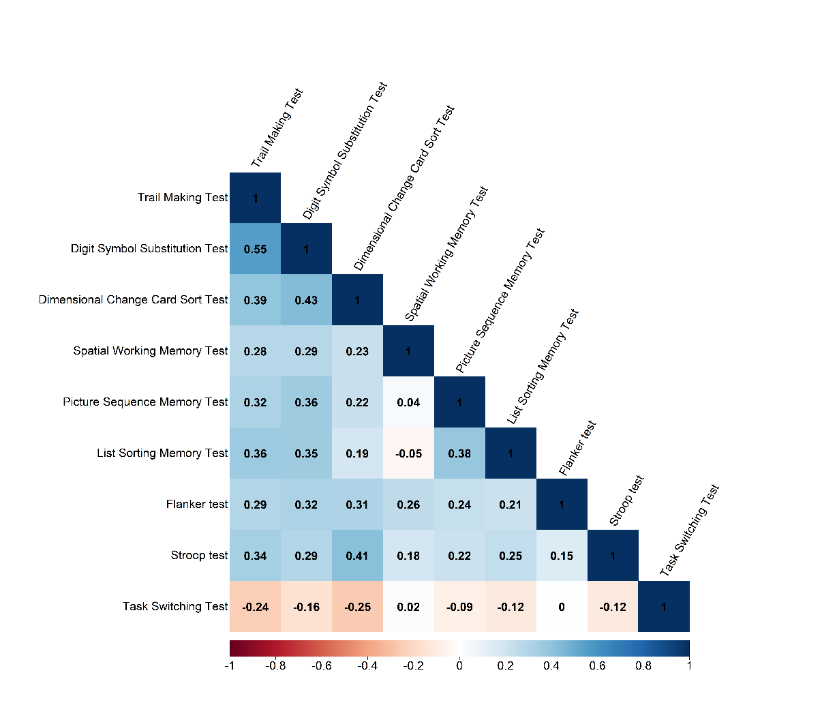

Figure S1. Pearson correlations between executive function measures. Color bar represents the strength of the correlation as r values. The darker blue the color, the stronger the positive correlation between variables, and the darker red the color, the stronger the negative correlation between variables. Blank spaces represent non – significant correlations. Trail Making Test, Dimensional Change Card Sort Test, Spatial Working Memory Test, Flanker test, Stroop test and Task Switching Test are reverted so that they can be interpreted in the same fashion as the rest of the outcomes (i.e., higher score indicates better performance).

*2.1.2.4 Bartlett’s test of sphericity.*

Bartlett’s test of sphericity was significant (c^2^(45) = 151.1341, p < 0.001). The KMO measure of sampling adequacy indicated that the strength of the relationship among variables was high (KMO = 0.62) and, thus, it was acceptable to proceed with subsequent analyses.

*2.1.2.5 CFA models for executive function score*

The fit of the three models for creating the executive function score is detailed below: (i) The three-order factor model did not satisfy the model fit criteria (CFI = 0.825, TLI= 0.738, RMSEA = 0.096, SRMR = 0.077, c^2^(24)= 0.009, p < 0.001). A negative variance on working memory was found. Due to this lower fit, a series of alternative CFA were examined, as described in the method section. (ii) The hypothesis of a model with only second-order factor did not satisfy the model fit criteria (CFI = 0.707, TLI= 0.590, RMSEA = 0.145, SRMR = 0.284, c^2^(15) = 0.000). It was also replicated in a second-order model, but did not fit neither (CFI = 0.885, TLI= 0.815, RMSEA = 0.097, SRMR = 0.071, c^2^(13) = 0.032).

(iii) Finally, the best-fit-model was composed of one first-order factor, SWMT, DCCST, DSST and TMT (CFI = 0.975, TLI= 0.925, RMSEA = 0.095, SRMR = 0.042, c^2^ (2) =0.165, p = 0.000). **Supplementary material 3, Figure S2** presents the latent factor constructs of the final model with standardized factor loadings on the paths. All loadings were statistically significant (all p<0.001), and all measures had loadings >0.45.

The post hoc power analysis revealed that a sample of N = 90 is associated with power larger than > 99.99% to reject a bad-fit model (df = 28) with an amount of misspecification corresponding to RMSEA = 0.08 and alpha = 0.05.

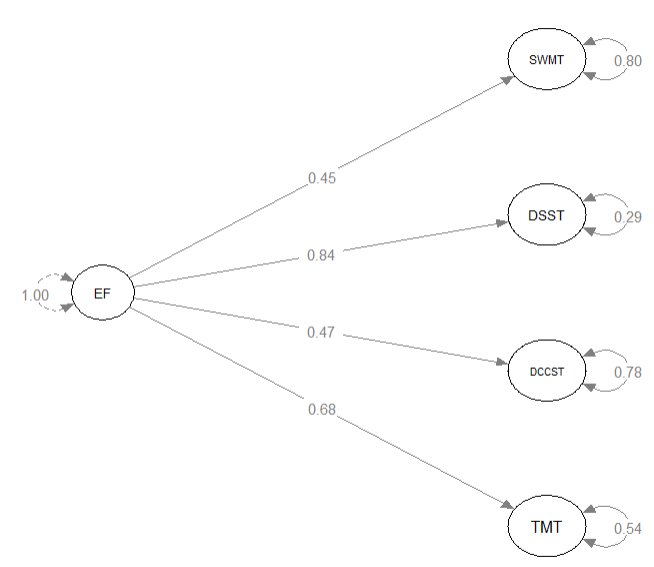

Figure S2. First-order factor analysis model. Factor loadings (numbers in the middle) represent the strength of the relationship between each test and the general executive function (EF) score. Higher loadings indicate a stronger association. The values next to each test represent the variance explained by the factor for each test. Abbreviations: TMT, Trail Making Test; DCCST, Dimensional Change Card Sort Test; SWMT, Spatial Working Memory Test; DSST, Digit Symbol Substitution Test.

*2.1.2.6 Summary of the executive function composite score*

The executive function composite score included the following sub-tests: SWM (43), DCCST (44), DSST (45) and TMT (46). The cognitive measure, outcome, range and mean are described in **Table S7**.

Table S7. Summary table with cognitive measure, outcome, range ad mean of the composite executive function score and the variables included in the score.

| Measure | Outcome (unit) | Range | Mean (SD) |
| --- | --- | --- | --- |
| Composite executive function score | Overall z-score | -3.38 - 1.55 | -0.01 (1.00) |
| *Sub-test included in the composite executive function score* | | | |
| Trail Making Test (TMT) | Interference Score  ( Time part B – Time part A) (sec) | -17.44 - 432.61 | 72.64 (74.1) |
| Digit Symbol Substitution Test (DSST) | Total correct score (n) | 18 - 74 | 44.54 (13.15) |
| Dimensional Change Card Sort Test (DCCST) | Inverse efficiency score (switch trials - high load) (rt/acc) | 0.01 - 0.07 | 0.01 (0.01) |
| Spatial Working Memory Test (SWMT) | Inverse efficiency score of all trials (rt/acc) | 8.01 - 48.33 | 15.29 (7.35) |

Abbreviations: acc, accuracy; n, number; sec, seconds; rt, reaction time. The Trail Making Test, Dimensional Change Card Sort Test, and Spatial Working Memory Test were reverted in the analyses, meaning that higher values indicate better performance. However, the data presented in the table are in their original (non-reverted) form. Baseline data are presented.

**3. Cognitive domains**

Cognitive domains were established by replicating a large-scale RCT with similar participant’s characteristics, cognitive tests and aims, but with a considerable larger sample size (n=648) to run a multiple-factor CFA model (35). Five cognitive domains were created: attentional/inhibitory control, episodic memory, processing speed, visuospatial processing and working memory. **Table S8** shows a summary of the cognitive measure included in each domain, along with their outcomes, ranges, and mean (SD) scores.

Table S8. Measure, outcome, range and mean of the domains included.

| Domain | Cognitive measure | Outcome (unit) | Range | Mean (SD) |
| --- | --- | --- | --- | --- |
| Attentional/inhibitory Control | Trail Making Test (TMT) | Time part B (sec) | 40.02 - 500.84 | 122.83 (85.6) |
|  | Dimensional Change Card Sort Test (DCCST) | Computed score* (n) | 2.13 - 8.56 | 6.96 (1.21) |
|  | Stroop Test | Incongruent raw score (rt) | 689.13 - 1364.34 | 970.84 (160.09) |
|  | Flanker Test | Computed score* (n) | 4.75 - 8.57 | 7.07 (0.82) |
| Episodic memory | MoCA Delayed Recall | Multiple choice cue (n) | 0.00 - 5.00 | 3.14 (1.25) |
|  | Picture Sequence Memory Test (PSMT) | Raw score (n) | 2.00 - 28.00 | 9.23 (5.53) |
|  | Rey Auditory Verbal Learning Test (RAVLT) | Total recall raw score- Sum of the 5 consecutive learning trials from List A (n) | 26.00 - 65.00 | 44.46 (8.36) |
|  |  | Total delayed recall correct raw score - Sum of words of List A after 20min (n) | 4.00 - 15.00 | 9.04 (2.54) |
|  |  | Recognition - Number of correct responses of the recognition trial - Number of intrusions false correct responses of the recognition trial) (n) | 8.00 - 15.00 | 14.03 (1.47) |
|  | Rey - Osterrieth Complex Figure Test (ROFT) | Raw score copy (n) | 11.50 - 36.00 | 32.66 (4.64) |
|  |  | Delayed raw score after 3 min period – Total of points (n) | 1.00 - 30.50 | 15.66 (7.19) |
| Processing Speed | Digit Symbol Substitution Test (DSST) | Total correct raw score (n) | 18.00 - 74.00 | 44.54 (13.15) |
|  | Trail Making Test (TMT) | Time part A (sec) | 21.28 - 218.63 | 50.19 (23.94) |
| Visuospatial processing | Wechsler Adult Intelligence Scale | Block design (acc) | 8.00 - 40.00 | 23.06 (7.18) |
|  |  | Matrix reasoning (acc) | 2.00 - 19.00 | 9.82 (4.67) |
|  | MoCA Clock Draw | Total Score (n) | 1.00 - 3.00 | 2.60 (0.6) |
| Working Memory | Spatial Working Memory Test (SWMT) | 3-item (acc) | 20.00 - 95.00 | 68.53 (15.74) |
|  |  | 4-item (acc) | 28.00 - 90.00 | 67.38 (13.62) |
|  | List Sorting Working Memory (LSWMT) | Total correct raw score (n) | 8.00 - 20.00 | 14.88 (2.17) |
|  | N-back | 2-back (acc) | 11.00 - 96.00 | 64.31 (22.51) |

Abbreviations: acc, accuracy; min,minutes; n, number; sec, seconds; rt, reaction time. Computed Score combines accuracy and reaction time (0-10). If accuracy ≤80%, score equals accuracy; if accuracy >80%, score is a combination. The Trail Making Test and Stroop Test variables were reverted in the analyses, meaning that higher values indicate better performance. However, the data presented in the table are in their original (non-reverted) form. Baseline data are presented.

**Supplementary material 4. Moderation outcomes**

**1. Moderators criteria**

To evaluate the moderating effects of individual characteristics, participants were categorized based on various key variables. The following sections outline the criteria and methods used to define each category.

***1.1 Sex***

Sex was categorized as male or female based on self-reported information regarding the sex assigned at birth.

***1.2 Age***

Age was dichotomized as youngers (<72 years old) or orders (≥72 years old), based on the median of study’s age range used for randomization (65-80 years).

***1.3 Educational level***

Educational level was grouped as low (≤12 years) or high (>12 years), based on the completion of high school or equivalent as a threshold.

***1.4 APOE4 carriership***

The genotyping of selected APOE SNPs (rs7412 and rs429358), which determine the presence of ε2, ε3, and ε4 alleles, was performed at GENYO using KASPar® assays (LGC Genomics, Hoddesdon, UK) following the manufacturer's protocol. Participants were classified as either APOE4 carriers (carrying at least one APOE 𝜀4 allele) or non-carriers.

***1.5 Amyloid burden***

Amyloid burden was assessed using positron emission tomography (PET) imaging and quantified with the Centiloid (CL) scale, which standardizes amyloid measurements across different methods. A cutoff of 12 CL was used to classify participants as amyloid-negative (<12 CL) or amyloid-positive (≥12 CL), based on established thresholds for identifying elevated amyloid burden.

***1.6 Comorbidities***

Participants were categorized into low (<3) or high (≥3–4) comorbidity group based on the total number of conditions reported in their self-reported medical history, including diagnoses or medications related to hypertension, diabetes, heart disease, obesity, or cholesterol.

***1.7 Baseline cognitive performance***

Baseline cognitive performance for EF and other cognitive domains, attentional/inhibitory control, episodic memory, processing speed, visuospatial processing and working memory, was assessed using pre-intervention data. Participants were categorized into two groups: low (<median) and high (≥median), based on the median scores calculated for each domain.

***1.8 Subjective cognitive decline***

The subjective cognitive decline was measured with the subjective cognitive decline scale (58), utilizing four questions related to self-experience. The questions were 1) Do you have more trouble remembering things that have happened recently? 2) Are you worse at remembering where belongings are kept? 3) Do you have trouble recalling conversations a few days later? 4) Do you have more trouble remembering appointments and social arrangements? All questions had the same options to answer with specific punctuation "No, not much worse (0 points)"; "A bit worse (1 point)"; "Yes, a lot worse (2 points)". Baseline cognitive decline was categorized as low (<median) or high (≥median) based on the sum of the four answers score from 0 to 8 points (59).

**Supplementary material 5. Physical condition parameters.**

**1. Overview of the muscular strength, physical function and cardiorespiratory fitness outcomes**

Additional outcomes related to physical condition parameters were divided into three categories: muscular strength, physical function, and cardiorespiratory fitness. Physical outcomes were assessed by using the Short Physical Performance Battery (SPPB), the Senior Fitness Test (SFT) and individual tests such as the handgrip test, the isokinetic tests or the 2-km walking test. A summary table detailing the measures, tests, and outcomes is presented in **Table S9**.

Table S9. Additional outcomes related to physical condition parameters: muscular strength, physical function, and cardiorespiratory fitness

| **Domain** | **Measure (battery)** | **Outcome (unit)** | **Range** | **Mean (SD)** |
| --- | --- | --- | --- | --- |
| Muscular strength | Arm Curl (SFT) | Repetitions (n/30sec) | 8.00 - 24.00 | 16.00 (3.77) |
| Muscular strength | 30-second sit-to-stand test (SFT) | Repetitions (n/30 sec) | 8.00 - 22.00 | 13.98 (2.97) |
| Muscular strength | 5-times sit-to-stand test (SFT) | Time (sec) | 5.16 - 16.76 | 9.20 (2.24) |
| Muscular strength | Handgrip test | Weight (Kg) | 12.65 - 48.65 | 28.03 (9.67) |
| Muscular strength | Isokinetic muscular strength; Elbow extension | Newton meters (Nm) | 16.00 - 247.50 | 89.90 (34.33) |
| Muscular strength | Isokinetic muscular strength: Knee extension | Newton meters (Nm) | 30.00 - 199.00 | 95.89 (33.02) |
| Physical function | 2-minute step test (SFT) | Repetitions (n/2min) | 38.00 - 170.00 | 70.83 (19.18) |
| Physical function | Up and go test (SFT) | Time (s) | 3.95 - 10.41 | 5.81 (1.13) |
| Cardiorespiratory fitness | 6-minute walk test (SFT) | Distance (m) | 318.80 - 693.70 | 477.84 (73.33) |
| Cardiorespiratory fitness | 2 km walking test | Time (min) | 15.27 - 35.67 | 23.29 (3.75) |
| Abbreviations: n, number; y, years; SD, standard deviation; sec, seconds; min, minutes; acc, accuracy; cm, centimeters; m, meters; N, Newtons; SFT, Senior Fitness Test; SPPB, Short Physical Performance Battery.  The 5-times sit-to-stand test (SFT), Up and go test (SFT) and 2 km walking test were reverted in the analyses, meaning that higher values indicate better performance. However, the data presented in the table are in their original (non-reverted) form. Baseline data are presented. | | | | |

**2. Muscular strength outcomes**

***2.1 Arm Curl***

The upper body muscular strength of participants was assessed using the arm curl test which is included in the SFT battery. Participants initiated the test from a seated position and, with a dumbbell (i.e., 5 pounds for females and 8 pounds for males), flexed and extended their dominant arm for a duration of 30 seconds upon the evaluator's signal. An explanatory demonstration of the test was given by the evaluator before its initiation. The test was carried out twice, and the highest score (repetitions) obtained was used as an upper body muscular strength indicator. Higher values indicate better performance.

***2.2 30-seconds sit-to-stand test***

The lower body strength of participants was evaluated through the 30-seconds sit-to-stand test, which is included in the SFT battery. Participants start the test from a seated position, crossing their hands over their chests, and were instructed to rise from the chair and sit down repeatedly for a continuous 30-second period. A demonstration of the test was provided by the evaluator prior to start. The test was conducted and recorded twice, but only the highest score (repetitions) was used as a lower body muscular strength indicator. Higher values indicate better performance.

***2.3 5-times sit-to-stand test***

The 5-times sit-to-stand test, included on the Short Physical Performance Battery, was employed to evaluate lower body strength. At the direction of the evaluator, participants, situated in a seated position with their hands crossed on their chests, were required to perform a sequence of standing up and sitting down from the chair for a total of five repetitions. Prior to the test, the evaluator elucidated the procedure through a demonstration. The test was executed twice, with both trials measured and recorded. The best time to completion (seconds) the five repetitions was used, and reverted (multiplied by -1) with higher values reflecting better performance.

***2.4 Handgrip strength test***

Isometric strength was measured using the handgrip test. Upon the evaluator's signal, participants grasped a dynamometer (Takei TKK 5401) tightly with their hand, exerting maximal force. The test was administered twice for each hand. Two scores of each hand were achieved in each test by the evaluator. Subsequently, the highest result for each hand was taken into account, and the average of the two hands was calculated to ascertain the isometric strength value in kg. Higher values indicate better performance

***2.5 Isokinetic muscular strength; Elbow extension***

Gymmex Iso-2 dynamometer (EASYTRCH s.r.l., Italy) was used to perform the Isokinetic Strength Test for upper limbs (elbow extension). The participant's initial position was as follows: they sat with their back fully against the back of the seat, holding the grip at the end with their non-performing hand. Their elbow was positioned at about a 130° angle to the horizontal, at chest height. Before starting the exercise, the participant received detailed instructions and completed a warm-up of 10 slow, controlled repetitions. For the exercise itself, the participant extended their elbow to a 180° position five times, applying maximum possible force each time. This was done sequentially with each arm. The score was obtained from the average of the five maximum-force knee extensions for both arms, expressed in Newton meters (Nm) as peak torque. Higher values indicate better performance.

***2.6 Isokinetic muscular strength ; Knee extension***

This test was performed using the same isokinetic device that was used for elbow extension, Gymmex Iso-2 dynamometer (EASYTRCH s.r.l., Italy). The participant’s initial position was as follows: the knee was bent at approximately a 90° angle to the horizontal, with the back fully supported against the seat, and both hands holding the grips at each end. Before starting the exercise, the participant received specific instructions and completed a warm-up. The exercise involved extending the knee to 180° a total of five times, applying maximum possible force each time. This was done sequentially with each leg. The score was obtained from the average of the five maximum-force knee extensions for both legs, expressed in Newton meters (Nm) as peak torque. Higher values indicate better performance

**3. Physical function outcomes**

***3.1 2-minute step test***

The 2-minute step test was employed to assess the physical function which is included in the SFT battery. Participants commenced the test by standing in front of a wall. The evaluator measured and marked the hip height on the wall. During the test, participants were required to alternately lift their knees above the marked height for a duration of two minutes. The evaluator provided a demonstration of the task and initiated the test by counting the number of times the participant's right knee exceeded the marked height on the wall. The number of repetitions in 2 minutes was used as indicator of this test and higher values indicate better performance.

***3.2 Up and go test***

The up and go test, which is included in the SFT battery, was conducted to assess walking speed, balance, and agility . Participants initiated the test from a seated position, while the evaluator positioned a cone at 2.5 meters in front of the participant. In response to the evaluator's cue, participants stood up, navigated around the cone, and returned to a seated position. The evaluator provided a demonstration to clarify the task and then timed the participant's test completion, emphasizing in the execution without resorting to running. The test was administered twice, and the shortest time recorded (In seconds) was used as the indicator of this test. The score was reverted (multiplied by -1) so higher values indicate better performance.

***3.3 Short-distance timed walk test***

The objective of this test was to assess participants' walking ability from the SFT battery. The test involved measuring the time taken to walk 4 meters in an unobstructed space. Participants were instructed to walk at a regular pace. Cones marked the start and end lines. Upon the evaluator's signal, participants start walking, and the evaluator measured the time. The test was repeated twice and the lower of the two trials (in seconds) was recorded as the result.

**4. Cardiorespiratory fitness outcomes**

***4.1 6-minute walk test***

The 6-minute walk test, from the SFT battery, measured cardiorespiratory fitness. Participants were instructed to walk continuously for a duration of six minutes along a predetermined course, maintaining a comfortable yet brisk pace without resorting to running. The evaluator set up a rectangular course using four cones (18.3m x 4.57m) positioned at each corner. After explaining the test, the evaluator initiated the timing using a stopwatch, and the same evaluator concluded the test at the end of the six-minute interval. Subsequently, the distance (In meters) covered by the participant was the indicator of cardiorespiratory fitness. Higher scores reflect better cardiorespiratory fitness

***4.2 2 km walking test***

The 2 km walking test involved participants walking a designated path. The evaluator arranged four cones (28m x 15m) at each corner to create a rectangle, along with an additional cone 22 meters from the starting cone to mark the end of the 2 km distance. The evaluator explained the test, which required participants to complete 23 full laps plus an additional 22 meters to reach the end cone. Upon initiating the timer using a stopwatch, the evaluator monitored the participant's progress. The same evaluator concluded the test upon the participant's arrival at the 2 km mark. The time taken (in minutes) by the participant was reverted (multiplied by -1) and recorded. Higher scores reflect better cardiorespiratory fitness.

**Supplementary material 6. Additional Tables and Figures**

| Table S10. Baseline descriptive characteristics of the AGUEDA sample. | | | |
| --- | --- | --- | --- |
|  | All | Exercise group | Control group |
|  | Mean ± SD | Mean ± SD | Mean ± SD |
| Overall characteristics | | | |
|  | 90 | 46 | 44 |
| Female, No. (%) | 52 (57.8) | 27 (58.7) | 25 (56.8) |
| Age at baseline, yr | 71.75 (3.96) | 71.90 (4.21) | 71.58 (3.73) |
| Education length, yr | 11.54 (4.90) | 11.17 (5.26) | 11.93 (4.53) |
| 12 yr of education or less, n (%) | 57 (63.3) | 32 (69.6) | 25 (56.8) |
| Height, cm | 160.44 (8.98) | 160.17 (8.87) | 160.72 (9.18) |
| Weight, kg | 73.50 (12.86) | 72.89 (13.41) | 74.14 (12.39) |
| Body-mass index, kg/m² | 28.50 (4.23) | 28.35 (4.43) | 28.67 (4.05) |
| More than 2 comorbidities , n (%) | 37 (41.1) | 15 (32.6) | 22 (50.0) |
| ApoE ε4 carrier, n (%) | 13 (14.8) | 7 (15.6) | 6 (14.0) |
| Amyloid positive, n (%) | 19 (21.1) | 8 (17.4) | 11 (25.0) |
| Subjective cognitive decline, score | 2.92 (1.97) | 2.74 (1.99) | 3.11 (1.94) |
| Cognitive status | | | |
| Telephone Interview of Cognitive Status, score | 33.91 (2.72) | 33.78 (2.65) | 34.05 (2.81) |
| Mini‑Mental State Examination , score | 28.92 (1.06) | 29.00 (0.97) | 28.84 (1.16) |
| Montreal Cognitive Assessment, score | 25.58 (2.14) | 25.28 (2.21) | 25.89 (2.05) |
| Cognitive function | | | |
| Executive function, z-score | -0.01 (1.00) | -0.03 (1.07) | 0.01 (0.94) |
| Episodic memory, z-score | 0.00 (1.01) | -0.01 (0.85) | 0.01 (1.15) |
| Processing speed, z-score | -0.01 (1.00) | 0.04 (0.85) | -0.06 (1.15) |
| Working memory, z-score | -0.02 (0.99) | 0.01 (1.05) | -0.04 (0.94) |
| Attentional/inhibitory control, z-score | -0.01 (1.00) | 0.04 (0.98) | -0.06 (1.03) |
| Visuospatial processing, z-score | -0.01 (1.00) | -0.09 (0.94) | 0.08 (1.06) |
| Muscular strength by field tests | | | |
| Arm curl, n | 16.00 (3.77) | 15.80 (3.59) | 16.20 (3.97) |
| 30-seconds sit-to-stand test, n | 13.98 (2.97) | 13.91 (3.03) | 14.05 (2.94) |
| 5-times sit-to-stand test, sec | -9.20 (2.24) | -9.22 (2.32) | -9.19 (2.17) |
| Handgrip strength, kg | 28.03 (9.67) | 27.18 (8.80) | 28.92 (10.53) |
| Muscular strength by Isokinetic tests | | | |
| Elbow extension, N/m | 89.90 (34.33) | 87.25 (35.98) | 92.67 (32.70) |
| Knee extension, N/m | 95.89 (33.02) | 93.08 (34.85) | 98.83 (31.13) |
| Physical function | | | |
| 2-Minute step, n | 70.83 (19.18) | 69.61 (21.72) | 72.11 (16.26) |
| Up and go test*, sec | -5.81 (1.13) | -5.72 (1.01) | -5.92 (1.24) |
| Cardiorespiratory fitness | | | |
| 6-Minute walk test, m | 477.84 (73.33) | 479.77 (73.58) | 475.82 (73.87) |
| 2 km walking test*, min | -23.29 (3.75) | -23.49 (4.13) | -23.08 (3.34) |
| Abbreviations: m, meters; n, number; N/m: Newton/meters, SD, standard deviation; sec, seconds; yr, years. *Reverted variable (*-1) for easy interpretation, higher score, the better performance. | | | |

| Table S11. Intention-to-treat effects of the 24-weeks resistance exercise intervention on executive function for subgroups. | | | |
| --- | --- | --- | --- |
|  | Executive function | | |
|  | All-Pre | Exercise-Post | Control-Post |
| Sex | | | |
| Male | 0.43 (0.18;0.68) | 0.54 (0.08;1) | 0.58 (0.09;1.07) |
| Female | -0.33 (-0.62;-0.04) | 0.29 (0.01;0.56) | 0.06 (-0.22;0.34) |
| Age (cut-off 72 years old) | | | |
| Youngers | 0.22 (-0.02;0.45) | 0.54 (0.18;0.9) | 0.55 (0.18;0.93) |
| Olders | -0.29 (-0.65;0.07) | 0.29 (-0.05;0.62) | -0.16 (-0.5;0.18) |
| Educational level (cut-off 12 years) | | | |
| High | 0.47 (0.24;0.71) | 0.67 (0.2;1.13) | 0.82 (0.39;1.26) |
| Low | -0.29 (-0.57;-0.01) | 0.27 (0;0.54) | -0.14 (-0.44;0.17) |
| APOE carrier | | | |
| Non-carrier | 0.02 (-0.21;0.25) | 0.5 (0.24;0.75) | 0.31 (0.05;0.58) |
| Carrier | -0.2 (-0.85;0.46) | -0.11 (-1.14;0.91) | 0.21 (-0.92;1.35) |
| Amyloid burden (cut-off 12 CL) | | | |
| Negative | -0.09 (-0.33;0.15) | 0.36 (0.06;0.65) | 0.27 (-0.05;0.6) |
| Positive | 0.29 (-0.12;0.71) | 0.51 (0.01;1.01) | 0.28 (-0.18;0.74) |
| Executive function (median) | | | |
| High | 0.7 (0.59;0.82) | 0.98 (0.77;1.18) | 1.01 (0.77;1.26) |
| Low | -0.73 (-1;-0.45) | -0.24 (-0.66;0.18) | -0.39 (-0.78;0) |
| Comorbidities (median) | | | |
| Low | -0.12 (-0.43;0.19) | 0.36 (0;0.73) | 0.22 (-0.18;0.62) |
| High | 0.14 (-0.11;0.4) | 0.36 (0;0.73) | 0.29 (-0.07;0.64) |
| Subjective cognitive decline (median) | | | |
| Low | -0.1 (-0.4;0.19) | 0.29 (-0.03;0.62) | 0.44 (0.08;0.81) |
| High | 0.15 (-0.11;0.41) | 0.57 (0.16;0.98) | 0.1 (-0.27;0.48) |
| Values indicate estimated marginal means at each time point (and 95% Confidence Intervals). | | | |

| Table S12. Intention-to-treat effects of the 24-weeks resistance exercise intervention on cognitive domains for subgroups. | | | | | | | | | | | | | | | |
| --- | --- | --- | --- | --- | --- | --- | --- | --- | --- | --- | --- | --- | --- | --- | --- |
|  | Attentional/inhibitory control | | | Episodic memory | | | Processing speed | | | Visuospatial processing | | | Working memory | | |
|  | All-Pre | Exercise-Post | Control-Post | All-Pre | Exercise-Post | Control-Post | All-Pre | Exercise-Post | Control-Post | All-Pre | Exercise-Post | Control-Post | All-Pre | Exercise-Post | Control-Post |
| Sex | | | | | | | | | | | | | | | |
| Male | 0.34 (0.05;0.64) | 0.98 (0.55;1.41) | 0.44 (-0.03;0.91) | 0.09 (-0.25;0.43) | 0.59 (0.02;1.17) | 0.3 (-0.32;0.92) | 0.4 (0.14;0.66) | 0.93 (0.53;1.33) | 0.68 (0.25;1.1) | 0.15 (-0.21;0.52) | 0.65 (0.16;1.14) | 0.6 (0.05;1.15) | 0.41 (0.1;0.71) | 0.71 (0.25;1.17) | 0.48 (-0.04;1) |
| Female | -0.27 (-0.55;0.01) | 0.47 (0.16;0.77) | 0.11 (-0.2;0.42) | -0.06 (-0.34;0.21) | 0.3 (-0.07;0.68) | -0.04 (-0.41;0.34) | -0.31 (-0.6;-0.02) | 0.05 (-0.28;0.37) | 0.3 (-0.02;0.63) | -0.13 (-0.38;0.12) | -0.2 (-0.53;0.13) | -0.22 (-0.56;0.12) | -0.32 (-0.59;-0.06) | 0.06 (-0.2;0.33) | 0.09 (-0.18;0.37) |
| Age (cut-off 72 years old) | | | | | | | | | | | | | | | |
| Youngers | 0.29 (0.05;0.53) | 0.98 (0.64;1.32) | 0.52 (0.16;0.88) | 0.24 (0.02;0.46) | 0.75 (0.32;1.18) | 0.37 (-0.09;0.84) | 0.15 (-0.08;0.39) | 0.63 (0.31;0.95) | 0.69 (0.36;1.02) | 0.16 (-0.13;0.44) | 0.34 (-0.04;0.72) | 0.27 (-0.14;0.68) | 0.11 (-0.16;0.38) | 0.4 (0.1;0.7) | 0.49 (0.17;0.81) |
| Olders | -0.39 (-0.73;-0.05) | 0.32 (-0.04;0.67) | -0.09 (-0.45;0.27) | -0.3 (-0.67;0.08) | -0.05 (-0.51;0.41) | -0.22 (-0.68;0.25) | -0.21 (-0.58;0.16) | 0.2 (-0.24;0.64) | 0.14 (-0.31;0.59) | -0.22 (-0.53;0.1) | -0.07 (-0.56;0.43) | -0.12 (-0.63;0.39) | -0.17 (-0.5;0.16) | 0.31 (-0.13;0.75) | -0.06 (-0.5;0.39) |
| Educational level (cut-off 12 years) | | | | | | | | | | | | | | | |
| High | 0.37 (0.15;0.58) | 1.04 (0.73;1.35) | 0.73 (0.45;1.02) | 0.45 (0.13;0.77) | 1.01 (0.48;1.53) | 0.81 (0.33;1.3) | 0.51 (0.28;0.73) | 1.06 (0.65;1.46) | 1.04 (0.68;1.41) | 0.46 (0.14;0.78) | 0.48 (-0.13;1.09) | 0.68 (0.14;1.21) | 0.46 (0.19;0.73) | 0.66 (0.18;1.13) | 0.79 (0.36;1.21) |
| Low | -0.23 (-0.52;0.07) | 0.51 (0.18;0.85) | -0.1 (-0.48;0.28) | -0.26 (-0.52;0) | 0.1 (-0.27;0.47) | -0.4 (-0.83;0.04) | -0.31 (-0.59;-0.03) | 0.1 (-0.22;0.42) | 0.08 (-0.28;0.43) | -0.28 (-0.53;-0.03) | -0.01 (-0.32;0.3) | -0.32 (-0.69;0.05) | -0.29 (-0.56;-0.02) | 0.21 (-0.06;0.47) | -0.18 (-0.5;0.13) |
| APOE carrier | | | | | | | | | | | | | | | |
| Non-carrier | 0.01 (-0.23;0.24) | 0.7 (0.4;1) | 0.26 (-0.04;0.57) | 0.04 (-0.2;0.28) | 0.49 (0.13;0.85) | 0.23 (-0.14;0.61) | 0.01 (-0.22;0.24) | 0.48 (0.17;0.78) | 0.52 (0.21;0.83) | 0.02 (-0.21;0.25) | 0.17 (-0.14;0.48) | 0.25 (-0.07;0.58) | 0.06 (-0.15;0.28) | 0.36 (0.1;0.62) | 0.43 (0.15;0.7) |
| Carrier | -0.08 (-0.7;0.53) | 0.52 (0.11;0.93) | 0.16 (-0.28;0.6) | -0.24 (-0.76;0.29) | 0.07 (-0.71;0.86) | -0.37 (-1.23;0.5) | -0.07 (-0.69;0.55) | 0.21 (-0.4;0.81) | 0.17 (-0.45;0.79) | -0.2 (-0.79;0.39) | 0.38 (-0.52;1.28) | -0.7 (-1.74;0.34) | -0.54 (-1.29;0.2) | 0.08 (-0.51;0.67) | -0.2 (-0.88;0.48) |
| Amyloid burden (cut-off 12 CL) | | | | | | | | | | | | | | | |
| Negative | -0.07 (-0.3;0.17) | 0.64 (0.34;0.94) | 0.15 (-0.18;0.47) | -0.04 (-0.29;0.21) | 0.37 (-0.01;0.74) | 0.08 (-0.32;0.48) | -0.02 (-0.26;0.21) | 0.43 (0.13;0.74) | 0.43 (0.1;0.76) | 0.03 (-0.21;0.28) | 0.16 (-0.17;0.48) | 0.3 (-0.07;0.66) | -0.08 (-0.33;0.17) | 0.35 (0.08;0.62) | 0.35 (0.05;0.65) |
| Positive | 0.2 (-0.28;0.68) | 0.86 (0.47;1.24) | 0.59 (0.22;0.96) | 0.15 (-0.26;0.55) | 0.75 (-0.27;1.76) | 0.17 (-0.62;0.97) | 0.05 (-0.43;0.53) | 0.38 (-0.15;0.9) | 0.57 (0.07;1.08) | -0.16 (-0.6;0.28) | 0.34 (-0.46;1.15) | -0.55 (-1.25;0.15) | 0.23 (-0.1;0.56) | 0.24 (-0.47;0.94) | 0.04 (-0.54;0.63) |
| Comorbidities (median) | | | | | | | | | | | | | | | |
| Low | -0.02 (-0.31;0.26) | 0.76 (0.42;1.1) | 0.13 (-0.24;0.49) | -0.08 (-0.36;0.2) | 0.51 (0.09;0.93) | -0.01 (-0.47;0.45) | -0.08 (-0.4;0.25) | 0.37 (0;0.74) | 0.41 (0.02;0.8) | 0.02 (-0.24;0.29) | 0.21 (-0.17;0.58) | 0.03 (-0.39;0.46) | -0.07 (-0.37;0.23) | 0.33 (0.02;0.63) | 0.25 (-0.1;0.59) |
| High | 0.01 (-0.31;0.33) | 0.53 (0.15;0.9) | 0.43 (0.07;0.79) | 0.12 (-0.21;0.45) | 0.14 (-0.36;0.63) | 0.33 (-0.16;0.82) | 0.09 (-0.14;0.32) | 0.5 (0.11;0.9) | 0.54 (0.17;0.92) | -0.06 (-0.41;0.3) | 0.15 (-0.38;0.67) | 0.15 (-0.35;0.64) | 0.06 (-0.23;0.35) | 0.36 (-0.11;0.84) | 0.28 (-0.17;0.73) |
| Subjective cognitive decline (median) | | | | | | | | | | | | | | | |
| Low | -0.08 (-0.37;0.21) | 0.64 (0.35;0.92) | 0.43 (0.12;0.74) | -0.14 (-0.42;0.15) | 0.37 (-0.01;0.76) | 0.26 (-0.18;0.69) | -0.03 (-0.31;0.26) | 0.37 (0.06;0.68) | 0.63 (0.29;0.97) | -0.15 (-0.42;0.12) | 0.15 (-0.19;0.49) | 0.32 (-0.09;0.72) | -0.09 (-0.34;0.17) | 0.31 (0.04;0.57) | 0.51 (0.21;0.81) |
| High | 0.11 (-0.19;0.4) | 0.77 (0.28;1.26) | 0.09 (-0.36;0.53) | 0.24 (-0.07;0.54) | 0.42 (-0.15;1) | 0.08 (-0.46;0.62) | 0.03 (-0.29;0.34) | 0.53 (0.06;1.01) | 0.28 (-0.17;0.73) | 0.23 (-0.1;0.57) | 0.15 (-0.41;0.72) | -0.07 (-0.58;0.43) | 0.1 (-0.28;0.49) | 0.43 (-0.07;0.94) | -0.03 (-0.48;0.42) |
| Baseline cognitive domain | | | | | | | | | | | | | | | |
| High | 0.79 (0.66;0.91) | 1.12 (0.79;1.45) | 0.93 (0.59;1.26) | 0.76 (0.59;0.93) | 1.14 (0.74;1.55) | 0.89 (0.49;1.29) | 0.72 (0.59;0.85) | 1.07 (0.83;1.32) | 1.28 (1.02;1.55) | 0.82 (0.68;0.96) | 0.72 (0.26;1.18) | 0.73 (0.26;1.2) | 0.79 (0.66;0.93) | 0.78 (0.41;1.15) | 0.74 (0.36;1.13) |
| Low | -0.81 (-1.03;-0.58) | 0.2 (-0.12;0.51) | -0.38 (-0.72;-0.05) | -0.76 (-0.98;-0.54) | -0.28 (-0.69;0.13) | -0.72 (-1.18;-0.26) | -0.74 (-1;-0.48) | -0.22 (-0.58;0.14) | -0.33 (-0.69;0.03) | -0.8 (-1;-0.6) | -0.28 (-0.58;0.03) | -0.6 (-0.95;-0.25) | -0.83 (-1.03;-0.62) | -0.11 (-0.39;0.18) | -0.24 (-0.56;0.07) |
| Values indicate estimated marginal means at each time point (and 95% Confidence Intervals). | | | | | | | | | | | | | | | |

| Table S13. Intention-to-treat effects of the 24-weeks resistance exercise intervention on physical parameters (raw data). | | |
| --- | --- | --- |
|  | Change from Baseline to 24-weeks | |
|  | Exercise | Control |
| Muscular strength by field tests | | |
| Arm curl, n | 18.49 (17.5;19.47) | 18.19 (17.15;19.22) |
| 30-seconds sit-to-stand test, n | 16.65 (15.59;17.7) | 15.49 (14.38;16.59) |
| 5-times sit-to-stand test, sec | -8.14 (-8.75;-7.54) | -8.6 (-9.23;-7.96) |
| Handgrip strength, kg | 27.88 (25.86;29.9) | 27.24 (25.2;29.28) |
| Muscular strength by Isokinetic tests | | |
| Elbow extension, N/m | 96.06 (88.56;103.57) | 92.42 (84.42;100.42) |
| Knee extension, N/m | 92.69 (86.09;99.29) | 84.31 (77.41;91.21) |
| Physical function | | |
| Two-Minute step, n | 88.1 (81.24;94.95) | 87.01 (79.67;94.35) |
| Up and go test, sec | -5.39 (-5.7;-5.09) | -5.45 (-5.77;-5.13) |
| Cardiorespiratory Fitness | | |
| 6-Minute walk test, m | 508.1 (489.4;526.79) | 498.02 (478.61;517.43) |
| 2 km walking test, min | -22.48 (-23.39;-21.57) | -23.34 (-24.29;-22.39) |
| Abbreviations: m, meters; n, number; N/m: Newton/meters, SD, standard deviation; sec, seconds; yr, years. Values indicated estimated marginal means (and 95% Confidence Intervals) after 24 weeks of exercise intervention. | | |

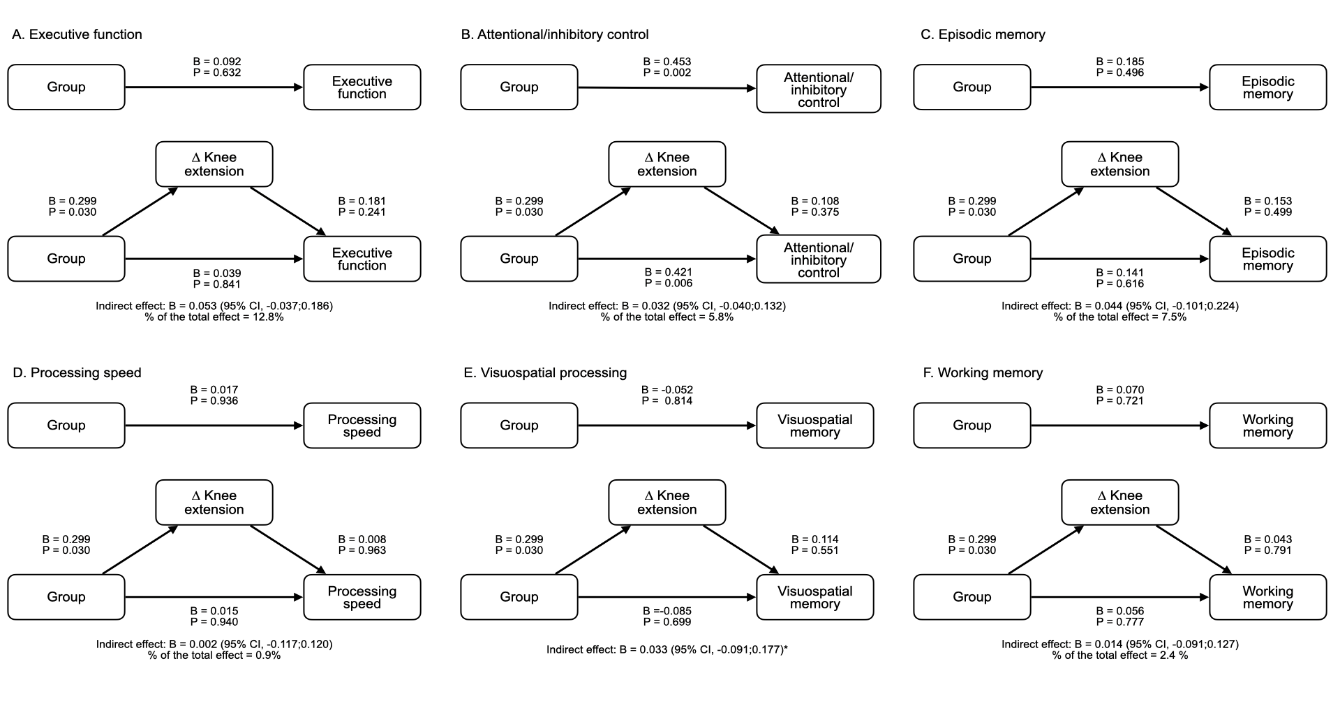

Figure S3. Mediation models of the Intervention effects knee extension on executive function and cognitive domains. *Negative % of mediation. △Knee extension represents the variable of change.

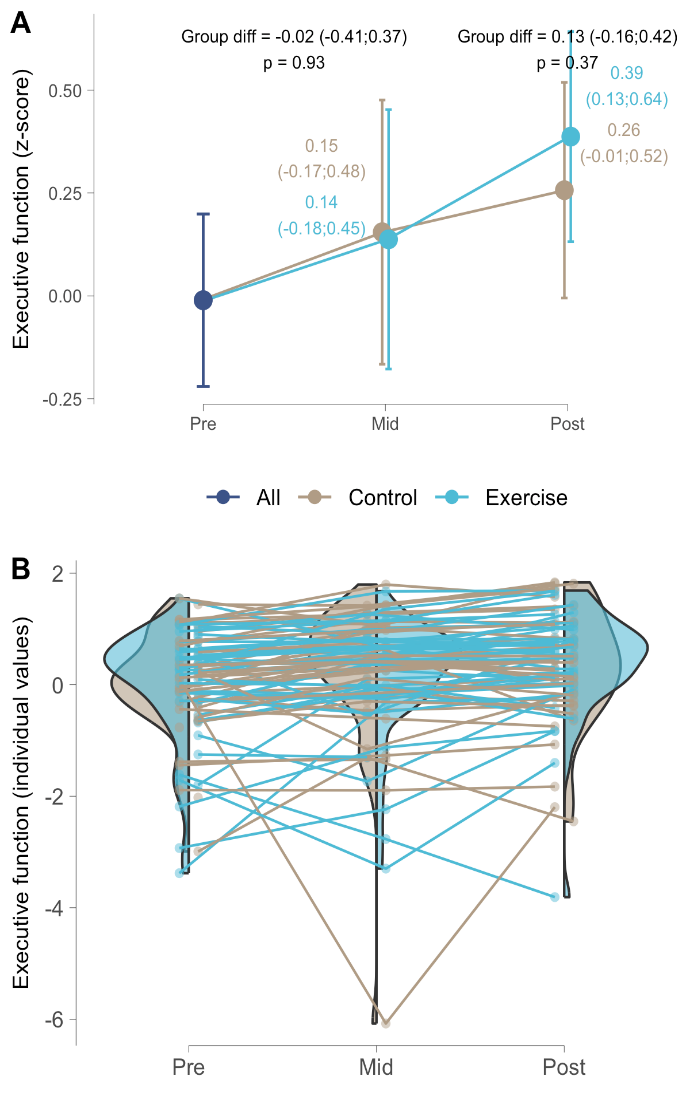

Figure S4. Intention-to-treat effects at 12 and 24 weeks of the resistance exercise intervention on executive function showing mean values (A) and individual values (B). A. Dots indicate estimated marginal means at each time point (and 95% Confidence Intervals). B. Dots indicate participant individual raw values.

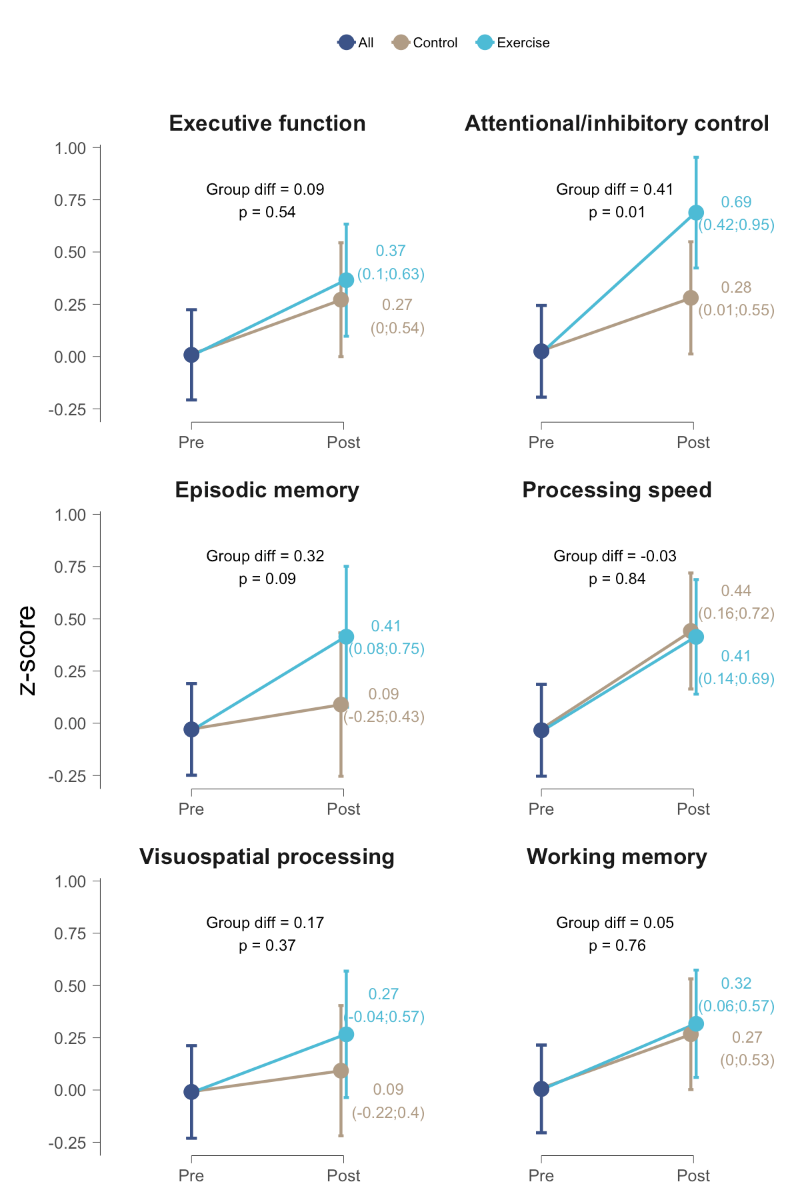

Figure S5. Per-protocol effects of the 24-week resistance exercise program on executive function and other cognitive domains. Dots indicate estimated marginal means at each time point (and 95% Confidence Intervals).

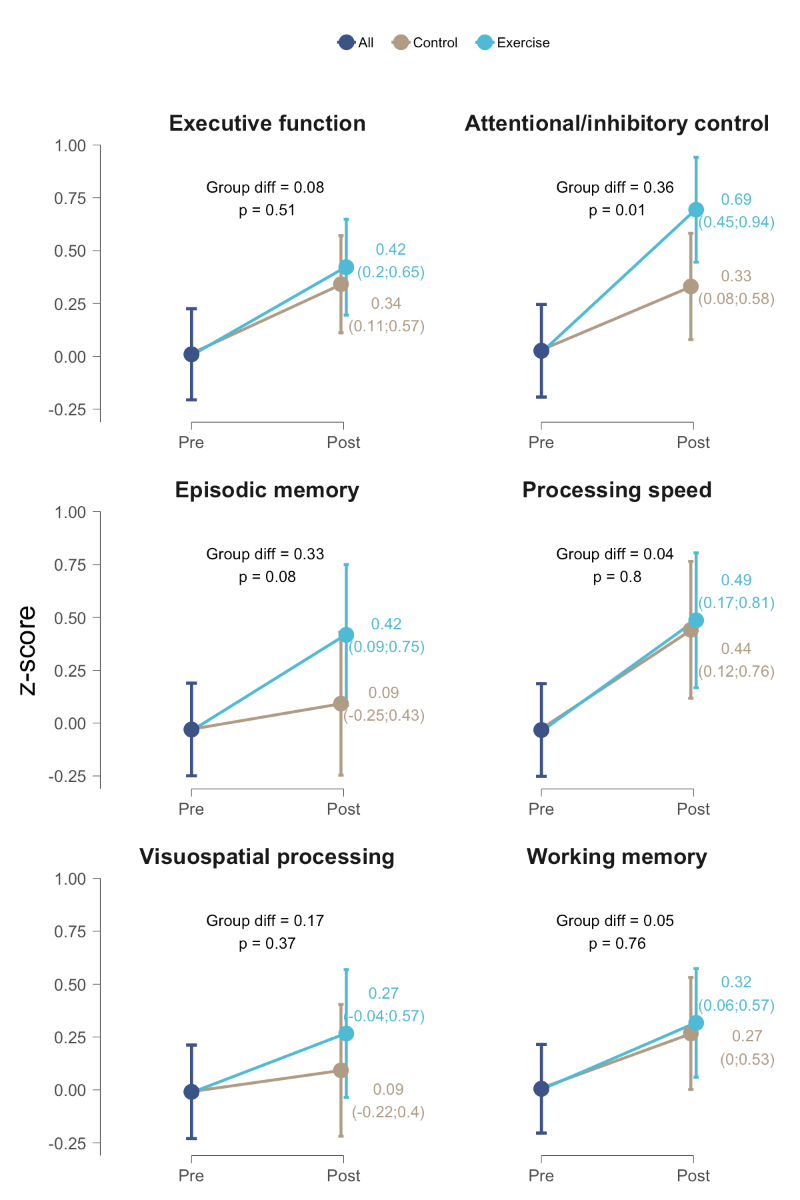

Figure S6. Per-protocol effects of the 24-week resistance exercise intervention on executive function and cognitive domains. Dots indicate estimated marginal means at each time point (and 95% Confidence Intervals). Winsorized values >4S

Table S14. Adverse events during 24-week period of exercise intervention.

| Adverse event # | Severity adverse event* | Group | Reason | Relationship with the Agueda Exercise intervention | Excluded from** | Action | Week of adverse event |
| --- | --- | --- | --- | --- | --- | --- | --- |
| 1 | Mild | Exercise group | Illness | No | Biological variables | Recommendations to visit the doctor | Week 17 |
| 2 | Mild | Exercise group | Knee pain | No | Lower body physical variables | Modifications and adaptations of the exercises | Week 22 |
| 3 | Moderate | Exercise group | Shoulder injury | No | Upper body physical variables | Modifications and adaptations of the exercises | Week 13 |
| 4 | Moderate | Exercise group | Shoulder injury | No | Upper body physical variables | Rehabilitation modifications and adaptations of the exercises | Week 2 |
| 5 | Moderate | Exercise group | Back pain | No | Upper and lower body physical variables | Modifications and adaptations of the exercises | Week 21 |
| 6 | Severe | Exercise group | Shoulder surgery | No | Upper body physical variables | Rehabilitation Modifications and adaptations of the exercises | Week 5 |
| 7 | Severe | Control group | Stage I of Cancer | No | All the variables | - | Week 12 |
| 8 | Severe | Control group | Stroke | No | All the variables | - | Week 18 |

*Mild, moderate or severe. **Those participants that had the adverse event closer to the 12 week or 24-week evaluations were excluded in sensitivity analyses
